# Nasal IgA wanes 9 months after hospitalisation with COVID-19 and is not induced by subsequent vaccination

**DOI:** 10.1101/2022.09.09.22279759

**Authors:** Felicity Liew, Shubha Talwar, Andy Cross, Brian J. Willett, Sam Scott, Nicola Logan, Matthew K. Siggins, Dawid Swieboda, Jasmin K. Sidhu, Claudia Efstathiou, Shona C. Moore, Chris Davis, Noura Mohamed, Jose Nunag, Clara King, A. A. Roger Thompson, Sarah L. Rowland-Jones, Annemarie B. Docherty, James D. Chalmers, Ling-Pei Ho, Alexander Horsley, Betty Raman, Krisnah Poinasamy, Michael Marks, Onn Min Kon, Luke Howard, Daniel G. Wootton, Susanna Dunachie, Jennifer K. Quint, Rachael A. Evans, Louise V. Wain, Sara Fontanella, Thushan I. de Silva, Antonia Ho, Ewen Harrison, J. Kenneth Baillie, Malcolm G. Semple, Christopher Brightling, Ryan S. Thwaites, Lance Turtle, Peter J.M. Openshaw, ISARIC4C Investigators and the PHOSP-COVID collaborative group

## Abstract

**Background:** Most studies of immunity to SARS-CoV-2 focus on circulating antibody, giving limited insights into mucosal defences that prevent viral replication and onward transmission. We studied nasal and plasma antibody responses one year after hospitalisation for COVID-19, including a period when SARS-CoV-2 vaccination was introduced.

**Methods:** Plasma and nasosorption samples were prospectively collected from 446 adults hospitalised for COVID-19 between February 2020 and March 2021 via the ISARIC4C and PHOSP-COVID consortia. IgA and IgG responses to NP and S of ancestral SARS-CoV-2, Delta and Omicron (BA.1) variants were measured by electrochemiluminescence and compared with plasma neutralisation data.

**Findings:** Strong and consistent nasal anti-NP and anti-S IgA responses were demonstrated, which remained elevated for nine months. Nasal and plasma anti-S IgG remained elevated for at least 12 months with high plasma neutralising titres against all variants. Of 180 with complete data, 160 were vaccinated between 6 and 12 months; coinciding with rises in nasal and plasma IgA and IgG anti-S titres for all SARS-CoV-2 variants, although the change in nasal IgA was minimal. Samples 12 months after admission showed no association between nasal IgA and plasma IgG responses, indicating that nasal IgA responses are distinct from those in plasma and minimally boosted by vaccination.

**Interpretation:** The decline in nasal IgA responses 9 months after infection and minimal impact of subsequent vaccination may explain the lack of long-lasting nasal defence against reinfection and the limited effects of vaccination on transmission. These findings highlight the need to develop vaccines that enhance nasal immunity.

**Research in context:** *Evidence before the study:* While systemic immunity to SARS-CoV-2 is important in preventing severe disease, mucosal immunity prevents viral replication at the point of entry and reduces onward transmission. We searched PubMed with search terms “mucosal”, “nasal”, “antibody”, “IgA”, “COVID-19”, “SARS-CoV-2”, “convalescent” and “vaccination” for studies published in English before 20^th^ July 2022, identifying three previous studies examining the durability of nasal responses that generally show nasal antibody to persist for 3 to 9 months. However, these studies were small or included individuals with mild COVID-19. One study of 107 care-home residents demonstrated increased salivary IgG (but not IgA) after two doses of mRNA vaccine, and another examined nasal antibody responses after infection and subsequent vaccination in 20 cases, demonstrating rises in both nasal IgA and IgG 7 to 10 days after vaccination.

*Added value of this study:* Studying 446 people hospitalised for COVID-19, we show durable nasal and plasma IgG responses to ancestral (B.1 lineage) SARS-CoV-2, Delta and Omicron (BA.1) variants up to 12 months after infection. Nasal antibody induced by infection with pre-Omicron variants, bind Omicron virus *in vitro* better than plasma antibody. Although nasal and plasma IgG responses were enhanced by vaccination, Omicron binding responses did not reach levels equivalent to responses for ancestral SARS-CoV-2. Using paired plasma and nasal samples collected approximately 12 months after infection, we show that nasal IgA declines and shows a minimal response to vaccination whilst plasma antibody responses to S antigen are well maintained and boosted by vaccination.

*Implications of all the available evidence:* After COVID-19 and subsequent vaccination, Omicron binding plasma and nasal antibody responses are only moderately enhanced, supporting the need for booster vaccinations to maintain immunity against SARS-CoV-2 variants. Notably, there is distinct compartmentalisation between nasal IgA and plasma IgA and IgG responses after vaccination. These findings highlight the need for vaccines that induce robust and durable mucosal immunity.

## Introduction

Intramuscular (i.m.) vaccines are remarkably effective in preventing severe COVID-19, their use being associated with declining hospitalisation.^1,2^ However, current vaccines provide only transient protection against respiratory viral replication, onward transmission and continuing emergence of variants. ^3–5^ By contrast, respiratory infection with SARS-CoV-2 induces mucosal immune defences that can inhibit viral replication and transmission, though the correlation between nasal and systemic immunity is inexact.^6,7^ To date, there have been few longitudinal studies of nasal antibody durability and those that exist give diverse results – suggesting that nasal antibody may persist for anywhere between 3 and 9 months.^8–10^ There is a clear need for additional studies of mucosal and systemic immunity in those recovered from severe disease.

Although i.m. vaccination transiently reduces transmission, vaccinees with breakthrough infections have peak nasopharyngeal viral loads similar to those in unvaccinated individuals. ^4,5^ Some studies have shown that viral loads decline more rapidly in vaccinees,^5^ but it is unclear whether this effect is mediated by passive transudation of plasma antibody into the mucosa, or whether vaccination can recall mucosal responses primed by infection (as observed after i.m. influenza vaccination following an intranasal (i.n.) priming).^11^ Serum IgA and IgG is mostly monomeric and produced in the bone marrow, whereas nasal IgA is polymeric and can be synthesized locally by mucosal plasma cells. It is polymeric nasal IgA that is critical for efficient neutralisation of virus in the upper respiratory tract, and so passive transudation of plasma antibody into the mucosa is unlikely to provide durable sterilizing immunity.^6^ Understanding whether i.m. vaccination after COVID-19 can recall nasal IgA responses is an important step towards developing vaccines which prevent infection and transmission.

During worldwide circulation of SARS-CoV-2, multiple successive variants have evolved, driven by enhancements in transmissibility as well as immune evasion. The Omicron subvariants appear less susceptible to vaccine-induced immunity and show high reinfection rates.^12,13^ It seems that immunity induced by successive infection and vaccination may provide superior protection against Omicron compared with either alone;^14,15^ and vaccination regimes which combine i.n and i.m. administration in mice induce enhanced mucosal protection against SARS-CoV-2 variants.^16^ This suggests that priming the nasal mucosa is required to induce effective local antibody responses that might provide enhanced immunity against current and future variants. However, the cross-reactivity of nasal antibody after infection with pre-Omicron virus is unknown.

We here report the results of a large multicentre longitudinal study of nasal and plasma antibody responses approximately a year after COVID-19, aiming to understand the longevity of nasal antibody responses after COVID-19 and the effect of subsequent vaccination. We demonstrate durable nasal and plasma IgG responses to ancestral (B.1 lineage) SARS-CoV-2, Delta and Omicron variant that are enhanced by i.m. vaccination. However, nasal IgA responses did not mirror those in plasma, waned after 9 months and were not substantially boosted by vaccination (Figure S1).

## Methodology

### Study design and participants

Clinical data, nasosorption and plasma samples were collected from hospitalised cases of COVID-19 within the ISARIC4C and PHOSP-COVID multicentre studies of UK adult patients (figure S2). ^17,18^

Adults hospitalised during the SARS-COV-2 pandemic were recruited into the International Severe Acute Respiratory and Emerging Infection Consortium (ISARIC) World Health Organization Clinical Characterisation Protocol UK (IRAS260007 and IRAS126600). Written informed consent was obtained from all patients. Ethical approval was given by the South Central–Oxford C Research Ethics Committee in England (reference: 13/SC/0149), Scotland A Research Ethics Committee (20/SS/0028) and World Health Organization Ethics Review Committee (RPC571 and RPC572l; 25 April 2013).

After hospital discharge patients >18 years old who had no co-morbidity resulting in a prognosis of less than 6 months, were recruited to the PHOSP-COVID study. Written informed consent was obtained from all patients. Ethical approvals for the PHOSP-COVID study were given by Leeds West Research Ethics Committee (20/YH/0225).

Samples and data were collected on day 1 to 9 of admission and/or at intervals during convalescence (approximately 1 to 14 months after discharge). Disease severity was classified according to the WHO Clinical Progression score.^19^

See supplementary materials for full methods.

### Procedures and immunoassays

Nasal samples were collected via nasosorption. Nasal and plasma IgA and IgG responses to Spike (S), Nucleocapsid (NP) and the Receptor-Binding-Domain of Spike (RBD) antigens of ancestral SARS-CoV-2 were measured using MSD (Mesoscale Diagnostics, Rockville, Maryland, USA) V-PLEX COVID-19 Coronavirus Panel 2 Kits. Antibody responses to RBD antigen of Delta and Omicron (BA.1) variants were measured using MSD V-PLEX SARS-CoV-2 panel 22. Nasal samples were diluted 1 in 50 and plasma 1 in 5000 prior to analysis. Nasosorption and plasma samples collected from 25 healthy participants prior to the emergence of SARS-CoV-2 were used as a control group.

Plasma neutralisation of ancestral SARS-CoV-2, Delta and Omicron (BA.1) variants was measured using a pseudotype neutralisation assay, as previously described at a dilution of 1 in 50.^20^ Samples with neutralising activity >90% were titrated to establish the titre resulting in 50% reduction in infectivity (PRNT_50_).

### Data analysis and Outcomes measured

Analyses were conducted using the Outbreak Data Analysis Platform (ODAP). Statistical analyses used R version 4.2.0. All tests were two-tailed and statistical significance was defined as a *p*-value<0.05 after adjustment for false discovery rate. Sample size calculations are detailed in supplementary materials.

Nasal antigen-specific IgA and IgG (AU/mL) was normalised to total isotype (pg/mL) accounting for concentration of sample obtained and matrix effects. Plasma and normalised nasal data were log_2_ transformed. The data were confirmed to be non-parametrically distributed using quantile Vs quantile plots. To understand the durability of antibody responses, comparisons between timepoints were made using the optimal pooled t-test, which performs well in non-parametric partially paired data.^21^ To estimate the effect of vaccination on antibody trajectories, a LOESS regression curve was fitted to data from repeated and cross-sectional samples taken from those who were known to be vaccinated.

To understand the relationship between compartments, disease severity and age, paired plasma and nasal responses taken from the same individuals were analysed in a correlation matrix measuring the Spearman rank correlation coefficient between variables. The variables in the correlogram were hierarchically clustered using Ward’s minimum variance. To further explore the relationship between nasal IgA and plasma IgA and IgG responses, unsupervised clustering was performed with Ward’s minimum variance and the results were visualised in a heatmap, which was subsequently annotated with age, disease severity and vaccination status to determine factors associated with cluster formation.

Control samples were used to define a nasal antibody threshold. The threshold was equivalent to the geometric mean titre (GMT) + 2 SD of controls and validated against standardized WHO BAU/mL thresholds converted into MSD AU/mL.^22^

## Results

A total of 446 adults, hospitalised between February 2020 and March 2021, were recruited and 569 plasma samples were collected, of which 338 represented samples taken from the same individual at sequential timepoints. In addition, 356 nasal samples were collected, of which 143 were taken from the same individual at sequential timepoints. 174 individuals had paired plasma and nasal samples taken at a given time point. Patient characteristics are shown in Table 1. The 6 and 12 month samples were collected between September 2020 and March 2022, covering the start of the UK vaccination campaign (figure S2).

**Table 1.** Summary of clinical and demographic data. Data are n (%) or median (IQR). Percentages were calculated after exclusion of missing data. Disease severity is classified according to the WHO Clinical Progression score: 3–4=no continuous supplemental oxygen needed; 5=continuous supplemental oxygen only; 6=continuous or bi-level positive airway pressure ventilation or high-flow nasal oxygen; 7–9=invasive mechanical ventilation or other organ support; and 10=did not survive. BMI=body-mass index.

| **Demographics (n=446)** |  |  | **Missing data** |
| --- | --- | --- | --- |
| **Age at admission, years** |  | 59 (51–67) | 28 (6.3) |
| **Sex at birth** | Female | 164 (39.1) | 27 (6.0) |
|  | Male | 255 (60.9) |  |
| **Ethnicity** | White | 259 (82.0) | 130 (29.0) |
|  | South Asian | 22 (6.9) |  |
|  | Black | 20 (6.3) |  |
|  | Mixed | 5 (1.6) |  |
|  | Other | 10 (3.2) |  |
| **Clinical characteristics (n=446)** |  |  | **Missing data** |
| **Disease severity** | WHO Class 3-4 | 60 (14.6) | 34 (7.6) |
|  | WHO Class 5 | 193 (46.8) |  |
|  | WHO Class 6 | 101 (24.5) |  |
|  | WHO Class 7-9 | 48(11.6) |  |
|  | WHO Class 10 | 10 (2.4) |  |
| **BMI ≥ 30** |  | 164 (62.1) | 183 (40.9) |
| **Co-morbidities** | None | 66 (20.8) | 129 (28.8) |
|  | 1 | 69 (21.8) |  |
|  | ≥2 | 183 (57.4) |  |
| **First vaccination received during the study** | Yes | 160 (89.9) | 266 (59.4) |
|  | No | 20 (11.1) |  |
| **Second vaccination received during the study** | Yes | 114 (65.5) | 272 (60.7) |
|  | No | 60 (34.5) |  |
| **Type of first vaccination** | Oxford/ AstraZeneca (ChAdOx1 nCoV-19) | 101 (64.7) | 290 (64.7) |
|  | Pfizer/Bio-N-Tec (BNT162b2) | 55 (35.3) |  |
|  | Moderna | 0 |  |
| **Type of second vaccination** | Oxford/ AstraZeneca (ChAdOx1 nCoV-19) | 65 (58.0) | 334 (74.6) |
|  | Pfizer/Bio-N-Tec (BNT162b2) | 46 (40.1) |  |
|  | Moderna | 1 (0.9) |  |

### Plasma antibody responses are more durable than nasal responses after COVID-19

Nasal anti-S and anti-NP IgA appeared within 4 weeks after symptom onset but waned after 9 months to levels equivalent to pre-pandemic controls (*p* < 0·0001) (figure 1). Anti-S IgG appeared within 14 days of symptom onset (*p*<0·0001) and rose 2181-fold after 9 months (p<0·0001) but unlike IgA responses, remained above pre-pandemic controls thereafter (p<0·0001) (figure 1 A–B). Both nasal IgA and IgG anti-S titres rose after 10 months, though the median change was only 1·46-fold in the case of IgA (*p*=0·011). Anti-NP IgA and IgG responses remained low after 9 months (*p*<0·0001) (figure 1 E–F).

**Figure 1.**
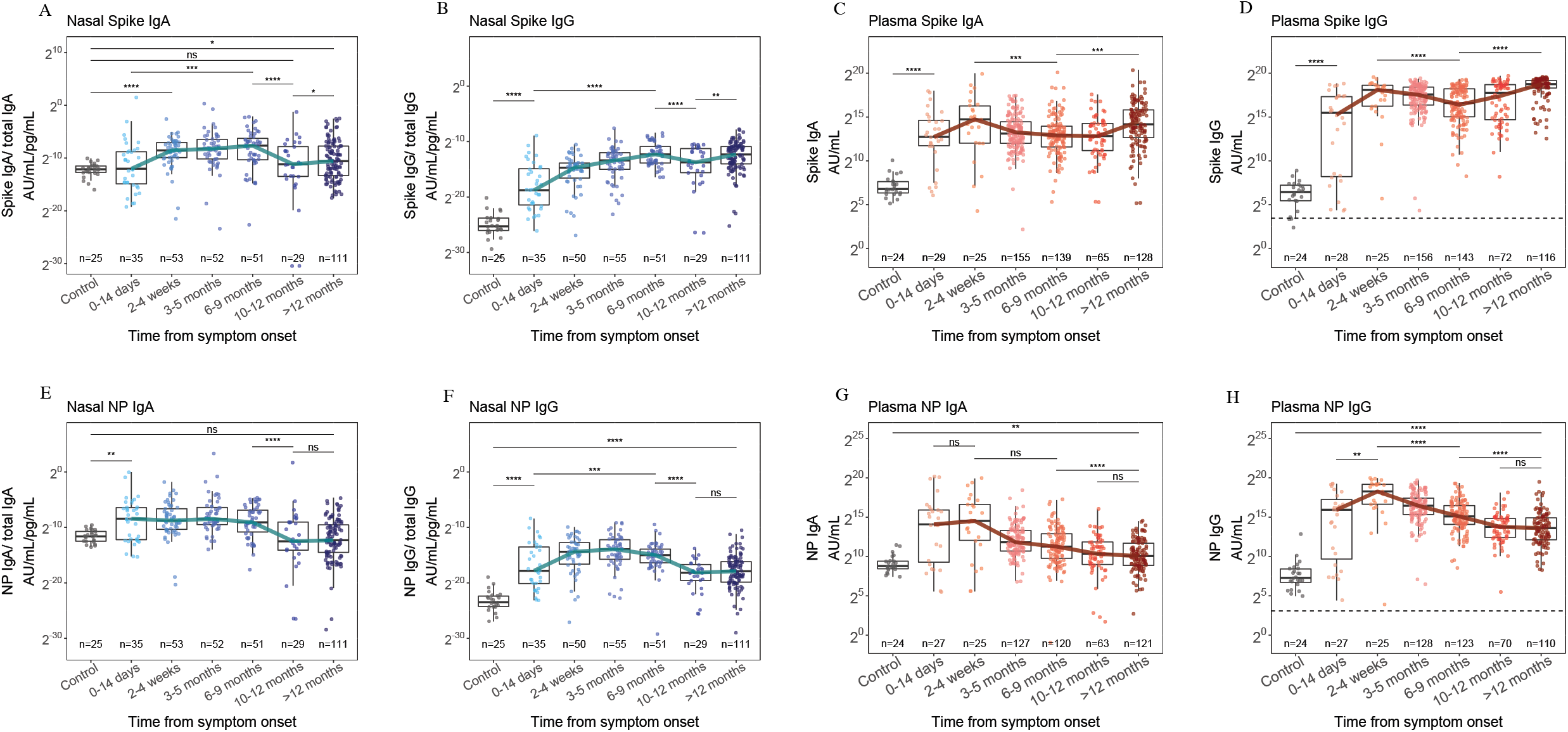
Nasal IgA (A), nasal IgG (B), plasma IgA (C) and plasma IgG (D) responses to S from ancestral SARS-CoV-2, 12 months after symptom onset and compared to pre-pandemic control samples (grey). Nasal IgA (E), nasal IgG (F), plasma IgA (G) and plasma IgG (H) responses to NP of ancestral SARS-CoV-2, 12 months after symptom onset and compared to pre-pandemic control samples. The blue and red lines indicate the trajectory of median titres across each timepoint. The horizontal dashed line indicates the WHO threshold for a seropositive titre. * = p<0·05, ** = p<0·01, *** = p<0·001, **** = p<0·0001.

Pre-pandemic controls allowed a threshold value for nasal antibody to be established, equivalent to the GMT+2SD (figure S3). Applying the same method to plasma samples, we found that the threshold value performed similarly to that of the WHO standards, confirming the validity of this method (figure S4). Using this threshold, we found that the nasal IgA GMT to S and NP fell below threshold after 9 months (Figure S3 A–B) whilst the nasal IgG GMT was durable and remained above threshold for both antigens at 12 months (Figure S3 F–G).

Plasma IgG anti-S and anti-NP responses developed within 14 days of symptom onset and remained elevated at 12 months (*p*<0·0001) (figure 1D and H). Notably, the trajectories of plasma IgA and IgG responses differed to that of nasal IgA. Whilst nasal responses peaked between 6 to 9 months for S and between 3 to 5 months for NP, plasma responses peaked within 4 weeks before waning (figure 1). Notably, plasma anti-NP responses plateaued after 10 months and all individuals were seropositive for both antigens at the final time point, indicating durable plasma responses after COVID-19 (figure 1D, G and H).

Only 2 of 446 individuals showed serological evidence of re-infection (whereby a rise in both anti-NP and anti-S IgG was observed between 103 and 308 days after infection for the first individual and between 238 and 463 days for the second). Furthermore, in 33 individuals where vaccination status was known and from whom samples were taken before and after vaccination anti-S titres rose (*p*<0·0001) whilst anti-NP titres declined (*p*=0·00019), as expected, indicating a low prevalence of re-infection in our cohort (figure S5 A–B). These data therefore demonstrate that nasal and plasma IgG responses are durable after COVID-19, whilst nasal IgA responses last only 9 months.

### Responses during vaccination campaign

Given the timing of vaccination in most of our cohort (median 20^th^ February 2021) and the timing of the 6 to 9 month visit (median 16^th^ March 2021), we reasoned that the increases in anti-S IgA and IgG seen in both nasal and plasma samples after 9 months were predominantly due to vaccination (figure 1 and S2). Of those with known vaccination status (n=180), 89% of individuals from whom plasma samples were collected and 95% of individuals from whom nasal samples were collected, received their first SARS-CoV-2 vaccination during the study. All vaccinations occurred between December 2020 and March 2022. Of these, 64.7% received ChAdOx1 nCoV-19 as the first dose (table 1). Since vaccines contain only S protein, NP responses are not induced by vaccination, and these responses remained low after 9 months (figure 1 E–H).

We confirmed the effect of vaccination by comparing S and NP antibody titres in individuals known to be vaccinated before and after their first vaccination (figure 2). Outliers who had samples taken >500 days after symptom onset were removed from this analysis to avoid modelling with insufficient data. Although the analysis was limited by the small number of nasal samples collected pre-vaccination (n=4), there were clear differences in the nasal IgA and IgG responses to S and NP after vaccination (figure 2 A– B). Although nasal anti-S IgA responses appeared elevated relative to anti-NP responses 100 days after vaccination, the difference in trajectories was small and the 95% Cis overlapped. By contrast, nasal IgG anti-S responses rose after vaccination and peaked approximately 150 days after vaccination, whilst the anti-NP trajectory declined. There was no overlap between the 95% confidence intervals (CI) of the regression curve for anti-S and anti-NP IgG responses after vaccination indicating distinct trajectories (figure 2B). Notably, the nasal IgG responses mirrored that of plasma IgA and IgG (figure 2 C–D). Thus, in keeping with the threshold analysis (figure S3), changes in nasal IgA titres after vaccination are minor compared to nasal and plasma IgG which are substantially boosted, suggesting that vaccination cannot fully recall mucosal antibody responses.

**Figure 2.**
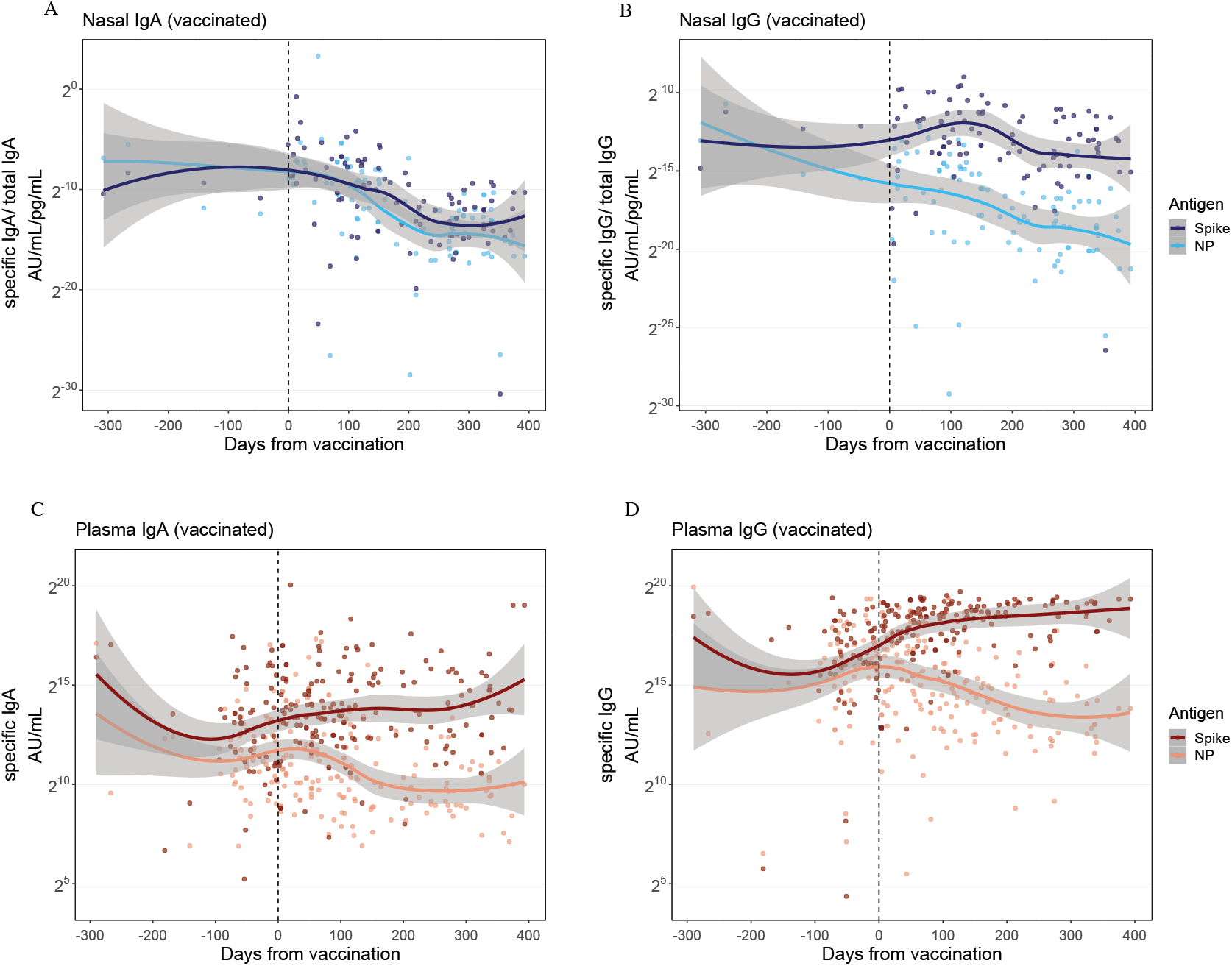
Trajectory of nasal IgA (A), nasal IgG (B), plasma IgA (C) and plasma IgG (F) anti-S and anti-NP responses before and after first vaccination. Trajectories have been modelled using a LOESS regression curve and 95% confidence intervals are shown in grey. The vertical dashed line indicates the time of first vaccination.

### Responses to Delta and Omicron (BA.1) variants

All participants were admitted to hospital prior to the emergence of Omicron variant and 71.1% (n=317) were admitted before 10^th^ May 2021 when Delta variant became the dominant strain in the UK.^4,12^ However, nasal IgA and IgG responses binding both Delta and Omicron RBD were present within 28 days of symptom onset and remained elevated for at least 9 months (figure 3 A–F). Nasal IgA binding Omicron appeared the most short-lived; the GMT only reached the threshold for positivity between 3 and 9 months (figure S3E). Furthermore, at its peak median titre, Omicron binding nasal IgA was only 10-fold above controls (*p*<0·0001), compared to nasal IgA binding ancestral SARS-CoV-2 RBD which was 28-fold higher (*p*<0·0001) (figure 3A and C). Plasma IgG responses to Delta and Omicron also developed within 14 days and were sustained for 12 months (figure 3 G–I).

**Figure 3.**
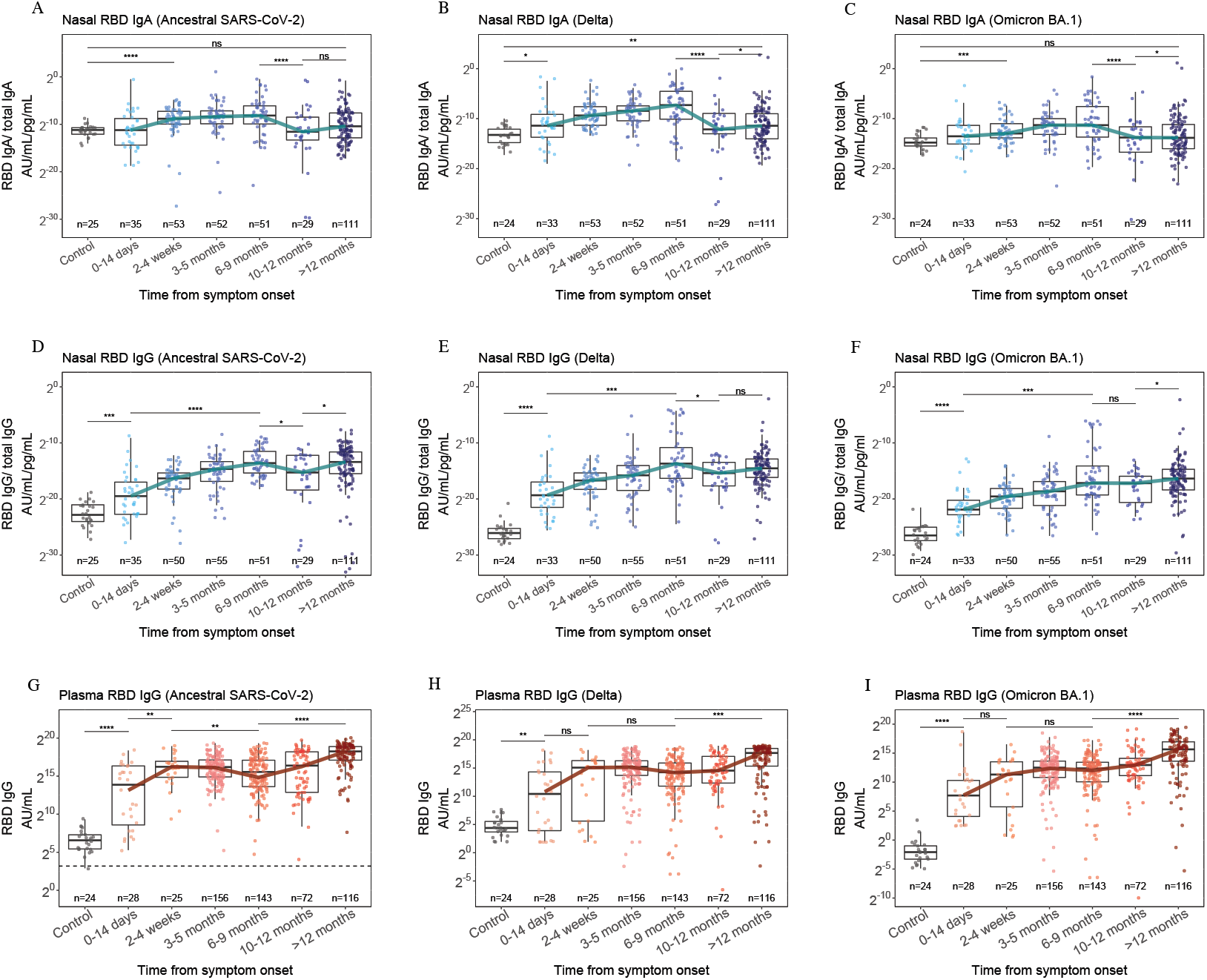
Nasal IgA (A–C), nasal IgG (D–G) and plasma IgG (G–I) responses to RBD of ancestral SARS-CoV-2, Delta, and Omicron (BA.1) variants 12 months after symptom onset compared to pre-pandemic control samples (grey). Nasal antibody titres have been normalised to total isotype content of sample. The blue and red lines indicate the trajectory of median titres across each timepoint. The horizontal dashed line indicates the WHO threshold for a seropositive titre. * = p<0·05, ** = p<0·01, *** = p<0·001, ****= p<0·0001.

To understand the degree of cross-reactivity between compartments we compared the ratio of antibody binding RBD of Omicron virus and ancestral SARS-CoV-2 (figure S6). There was no difference in the median ratio between nasal IgA (0·10) and nasal IgG (0·12, *p*=0·67). However, the nasal IgG ratio was higher than that of plasma IgG (0·09, *p*=0·020) and the nasal IgA ratio was higher than that of plasma IgA (0·08, *p*=0·00059). These data indicate that infection with pre-Omicron

SARS-CoV-2 can induce nasal and plasma antibody that binds Omicron RBD, and that nasal antibody may have greater cross-binding potential. However, despite this, Omicron-binding nasal IgA is slow to reach positive levels and is transiently maintained.

### Responses to Delta and Omicron (BA.1) variant after vaccination

The nasal IgA trajectory did not appear substantially different after vaccination (figure S7A) though a small rise in the Omicron- and Delta-binding nasal IgA GMT was seen between 10 and 12 months, when most individuals with known vaccination status had been vaccinated (figure 3 and S3). However, this difference was small and did not reach the positive threshold. Nasal IgG responses to Omicron and Delta variant rose after vaccination (figure S7B), although Omicron-binding responses did not reach the level of those to Delta and ancestral SARS-CoV-2 despite vaccination.

Plasma IgG responses to Delta and Omicron variants also rose after vaccination (figure 7 C–D). To study the effect of vaccination specifically, we identified 33 individuals from whom pre- and post-vaccination plasma samples were collected; these were taken at a median of 54 days (IQR 25·6–68·8) before the first vaccination dose and 176 days (IQR 113–212) after (figure S5). Vaccination substantially boosted Omicron-binding plasma IgG in these individuals, which rose 8·7-fold (p<0·0001). However, no significant difference in Delta-binding titres was seen, as antibody was boosted in some individuals but declined in others (figure S5D). This pattern may relate to a rapid rise and wane of vaccine-boosted antibody, given that samples were taken a median of 168 days after first vaccination and Delta-binding plasma IgG responses were observed to wane approximately 75 days from first vaccination (figure S7D). The subsequent rise in Delta-binding plasma IgG 150 days after vaccination likely results from individuals receiving their second vaccination dose during the study (table 1). These data suggest that vaccination can boost Omicron- and Delta-binding nasal and plasma IgG but enhancement of Delta responses may be short-lived after one vaccine dose. Meanwhile, Omicron- and Delta-binding nasal IgA responses are not significantly affected by vaccination.

### Plasma neutralising antibody to SARS-CoV-2 variants

Plasma neutralising titres against ancestral, Delta and Omicron variants of SARS-CoV-2 remained substantially elevated compared with controls between 3 and 12 months (figure S8). However, neutralising titres against Omicron were generally lower: at 10 to 12 months, 76·2% had neutralising antibody against Omicron, compared to 92·5% against ancestral SARS-CoV-2. Neutralising titres against all three variants were boosted during the vaccination campaign (*p<*0·0001) indicating that i.m. vaccination after COVID-19 can enhance neutralising antibody levels to homologous and heterologous variants.

As expected, neutralising antibody titres correlated with plasma RBD (*R*=0·82, *p*<0·0001) and S IgG (*R*=0·81, *p*<0·0001) (figure S9). Notably, plasma neutralising antibody correlated with nasal anti-RBD IgG (*R* =0·59, *p*<0·0001) and anti-S IgG (*R*=0·56, *p*<0·0001) but not nasal IgA (anti-RBD *R* = 0·1, *p*=0·39). This finding, alongside the boosting of nasal IgG after vaccination indicate that nasal IgG responses reflect that of plasma, whilst the nasal IgA response is distinct and compartmentalised (figure 2 and S9).

### Discordance between plasma and nasal antibody responses

To characterise the relationship between compartments, paired nasal and plasma samples from 175 individuals were examined. Samples were divided into those taken at approximately 6 months (3–9 months) and 12 months (>10–12 months) after infection (figure 4). At 6 months nasal anti-S IgA responses correlated strongly with nasal anti-NP IgA responses (*R*=0·71, *p*<0·0001) but showed a weaker association with nasal anti-S IgG (*R* =0·57, *p*<0·0001) and plasma anti-S IgA responses (*R* =0·50, *p*<0·0001) (figure 4A). There was no association between nasal IgA responses and plasma IgG response to either S (*p*=0·38) or NP (*p*=0·56). Nasal anti-NP IgA did not correlate with either nasal or plasma anti-NP IgG and correlated weakly with plasma anti-NP IgA (*R*=0·40, *p*=0·0021). Nasal IgG responses correlated with plasma IgG responses to the corresponding antigen, (anti-S *R*=0·47, *p*<0·0001 and anti-NP *R*=0·6, *p*<0·0001). A similar degree of compartmentalisation was observed at 12 months when the association between nasal and plasma anti-S IgA was even weaker (*R* =0·35, *p*<0·0001) and the association between nasal and plasma anti-S IgG was marginally stronger *(R*=0·51, p<0·0001) (figure 4B). Age and disease severity showed no association with nasal responses at both time points.

**Figure 4.**
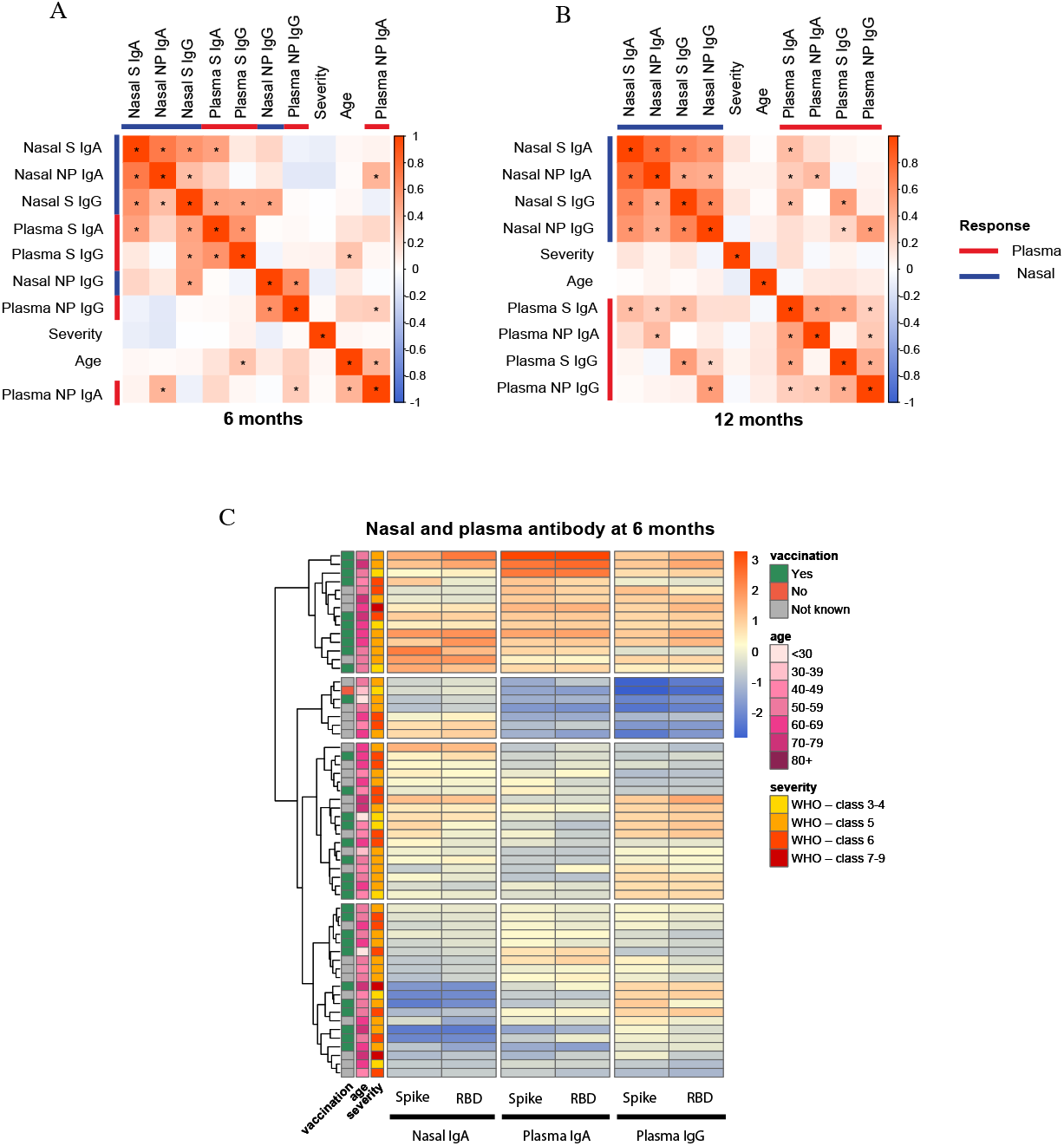
Correlogram of nasal and plasma IgA and IgG responses to S and NP, disease severity and age at 6 months, when 27 of 31 individuals with known vaccination status had received their first vaccination (A) and 12 months, when 58 of 63 individuals with known vaccination status had received both vaccinations (B). All statistically significant correlations are denoted with *. The variables were hierarchically clustered. Heatmap (C) of nasal IgA, plasma IgA and plasma IgG responses to S and RBD at 6–9 months. Rows are annotated with vaccination status, age and disease severity according to the WHO clinical progression score: 3–4 = no continuous supplemental oxygen needed; 5=continuous supplemental oxygen only; 6=continuous/bi-level positive airway pressure ventilation or high-flow nasal oxygen; 7–9=invasive mechanical ventilation or other organ support.

We considered the role of vaccination in driving the compartmentalisation between nasal IgA and plasma responses. At 6 months, 27 of 31 individuals with known vaccination status had received their first vaccination and 10 had received both doses. The median time from first vaccination was 81 days (IQR 20–105). Meanwhile at 12 months, 58 of 63 individuals with known vaccination status had received both vaccinations and the median time from second vaccination was 171 days (IQR 103– 246). Thus, we reasoned that the increased compartmentalisation between these time points may result from vaccination; whereby plasma responses are enhanced but nasal IgA is minimally affected.

To explore the relationship between nasal IgA and plasma antibody responses after first vaccination, we performed hierarchical clustering of anti-S/RBD responses from paired samples collected at 6 months (median 81 days after vaccination). Compartmentalisation of nasal IgA from plasma responses was observed with 4 distinct clusters forming (figure 4C). The first cluster exhibited patients with robust nasal IgA and plasma responses. Patients with the weakest plasma IgA and IgG responses were present in cluster 2 whilst patients with the weakest nasal IgA responses were in cluster 4. Although not statistically significant, there was a tendency towards more recent vaccination in cluster 1 compared with cluster 4 (figure S10A). The date of vaccination was not available for any members of cluster 2. Cluster 1 also contained a higher proportion of individuals receiving BNT162b2 vaccination (44%) compared with cluster 4 (27%), although the difference in proportions did not reach statistical significance due to the number of participants with complete vaccination data (figure S10B). There was no association between disease severity or age and cluster membership (figure 4C). Thus, we concluded that the clusters resulted from transient boosting of nasal IgA responses after recent vaccination, with divergence between the nasal IgA and plasma responses with increasing time from vaccination. Given the insubstantial and transient effect of vaccination on nasal IgA responses relative to plasma responses, we suggest that i.m. vaccination after COVID-19 is unable to adequately recall mucosal responses.

## Discussion

We demonstrate durable nasal and plasma IgG responses to ancestral (B.1 lineage), Delta and Omicron variants of SARS-CoV-2 in 446 adults hospitalised with COVID-19, who were infected with pre-Omicron virus and the majority of whom were subsequently vaccinated. However, we found that nasal virus-specific IgA levels fell back to pre-COVID levels after 9 months and Omicron-binding nasal responses were particularly short-lived. Our results reveal that nasal IgA responses are compartmentalised from systemic responses after vaccination, which boosted nasal and plasma IgG but not nasal IgA.

The durability of nasal antibody responses has hitherto been unclear. Whilst a Dutch study of healthcare workers found that nasal antibody lasted 9 months after mild infection, others demonstrated rapid waning after 3 months.^8,9^ Neither study examined a large cohort of hospitalised patients, and our findings confirm that COVID-19 can induce durable mucosal immunity. We also found that disease severity and age did not impact the longevity of the nasal responses in keeping with a recent study of 26 unvaccinated individuals.^10^

By calibrating nasal antibody levels with pre-COVID samples, we demonstrate that on average, nasal IgA responses disappear after 9 months and Omicron-binding IgA is particularly short-lived. Nasal IgA is the most abundant mucosal antibody and provides an important first-line defence against respiratory infection. The importance of nasal IgA in mediating immunity to SARS-CoV-2 is highlighted by a recent study where nasal IgA but not IgG correlates with nasal neutralisation after COVID-19.^10^ The short-lived nasal IgA response demonstrated here may explain the high rates of infection with Omicron variant, despite vaccination, and are in-keeping with real-world data reported in preprint, showing that infection with pre-Omicron virus has minimal influence on the risk of Omicron infection at 15 months.^13,23^

Whilst we found that i.m. vaccination can boost nasal IgG, it had limited effects on IgA, in keeping with a previous study of salivary antibody in 107 care home residents.^24^ We demonstrated correlations between nasal IgG, plasma IgG and plasma neutralisation, whilst nasal IgA responses were compartmentalised, suggesting that the rise in nasal IgG after vaccination could derive from plasma. Notably, we demonstrate that those exhibiting more robust nasal IgA responses had been recently vaccinated and a higher proportion had been vaccinated with BNT162b2 vaccine. Although this analysis was limited by small sample size, our findings suggest that vaccination only transiently boosts nasal IgA, and the type of vaccination received may influence the strength of response. mRNA vaccines tend to induce stronger circulating antibody responses than those using adenoviral vectors, and this may also apply to nasal responses.^25,26^ Taken together, these findings suggest that i.m. vaccination after COVID-19 cannot recall mucosal responses.

The concept of independent mucosal and systemic immunity is supported by recent studies showing that SARS-CoV-2 naïve individuals (whose mucosa have not been primed) do not produce nasal antibody after i.m. vaccination, highlighting that an independent response must occur at mucosal sites.^9,27^ Moreover, previous work has demonstrated that transudation of plasma antibody makes minimal contribution to total antibody concentrations in the mucosa, even in cases of paraproteinaemia where plasma concentrations are extremely high.^28^ This would explain why i.m. vaccination has had only transient effects on transmission,^4^ since the enhancement of nasal IgG that we observe, while measurable, is unlikely to have a considerable effect on mucosal susceptibility to infection. Future vaccines will need to substantially boost nasal IgA if they are to fully prevent infection and transmission. To date, intranasal and aerosolized vaccines have shown the most promise in doing so.^27,29,30^ It is therefore essential to prioritise development of mucosal vaccines which can provide better protection against respiratory infections.

### Study limitations

Although 338 individuals had samples taken from more than 1 time point after hospital discharge, given the circumstances and scale of this study we were not able to collect longitudinal samples from each participant. However, given that most individuals follow similar antibody kinetics, where longitudinal samples were missing, data were compared to cross-sectional samples taken from individuals in the acute and early convalescent phase of illness.

We did not have vaccination data for all cases, preventing direct comparison of pre- and post-vaccination nasal antibody titres. However, we demonstrated differences in nasal anti-S and anti-NP responses during the period of vaccination, enabling inferences to be drawn. Notably we estimated a peak of nasal anti-S IgG titres 150 days after vaccination which is considerably slower than peak circulating antibody responses after vaccination (28–42 days).^31^ Future studies using longitudinal data collected at fixed intervals before and after vaccination will better capture the peak of nasal antibody titres after i.m. vaccination.

## Conclusions

This is the first study to demonstrate durable but compartmentalised nasal IgA and plasma antibody responses to SARS-CoV-2 after infection and subsequent vaccination. We show enhancement of nasal and plasma IgG responses to ancestral SARS-CoV-2, Delta and Omicron variants after vaccination. However, nasal IgA responses, especially those to Omicron, are more short-lived and are not substantially affected by vaccination. Our results explain the lack of long-term sterilising immunity after previous infection and/or vaccination and highlight the need for mucosal vaccines that target nasal IgA responses. By enhancing nasal antibody responses, mucosal vaccines might prevent infection and transmission more effectively, enabling greater control of the pandemic and limiting the emergence of variants.

## Supporting information

The full list of authors within these groups is available in the supplementary materials.

The full list of authors within these groups is available in the supplementary materials.

## Data Availability

The ISARIC4C protocol, data sharing and publication policy are available at https://isaric4c.net. ISARIC4C Independent Data and Material Access Committee welcomes applications for access to data and materials (https://isaric4c.net). The PHOSP-COVID protocol, consent form, definition and derivation of clinical characteristics and outcomes, training materials, regulatory documents, information about requests for data access, and other relevant study materials are available online: https://phosp.org/resource/. Access to these materials can be granted by contacting the PHOSP-COVID collaborative group.

https://phosp.org/resource/.

https://isaric4c.net.

## Acknowledgements

This research used data assets made available by Outbreak Data Analysis Platform (ODAP) as part of the Data and Connectivity National Core Study, led by Health Data Research UK in partnership with the Office for National Statistics and funded by UK Research and Innovation (grant ref MC_PC_20058).

This work is supported by the following grants: The PHOSP-COVD study is jointly funded by UK Research and Innovation and National Institute of Health and Care Research (grant references: MR/V027859/1 and COV0319). ISARIC4C is supported by grants from the National Institute for Health and Care Research (award CO-CIN-01) and the Medical Research Council (grant MC_PC_19059) Liverpool Experimental Cancer Medicine Centre provided infrastructure support for this research (grant reference: C18616/A25153). Other grants which have supported this work include: the UK Coronavirus Immunology Consortium [funder reference:1257927], the Imperial Biomedical Research Centre (NIHR Imperial BRC, grant IS-BRC-1215-20013), the Health Protection Research Unit (HPRU) in Respiratory Infections at Imperial College London and NIHR HPRU in Emerging and Zoonotic Infections at University of Liverpool, both in partnership with Public Health England, [NIHR award 200907], Wellcome Trust and Department for International Development [215091/Z/18/Z], Health Data Research UK (HDR UK) [grant code: 2021.0155], Medical Research Council [grant code: MC_UU_12014/12], and NIHR Clinical Research Network for providing infrastructure support for this research.

FL is supported by an MRC clinical training fellowship [award MR/W000970/1]. LPH is supported by Oxford NIHR Biomedical Research Centre. AART is supported by a BHF Intermediate Clinical Fellowship (FS/18/13/33281). RAE holds a NIHR Clinician Scientist Fellowship (CS-2016-16-020). SD is funded by an NIHR Global Research Professorship [NIHR300791]. DW is supported by an NIHR Advanced Fellowship. PJMO is supported by a NIHR Senior Investigator Award [award 201385]. LT is supported by the Wellcome Trust [clinical career development fellowship grant number 205228/Z/16/Z], the Centre of Excellence in Infectious Diseases Research (CEIDR) and the Alder Hey Charity.

The funders were not involved in the study design, interpretation of data or writing of this manuscript. The views expressed are those of the authors and not necessarily those of the DHSC, DID, NIHR, MRC, the Wellcome Trust, UK-HAS, the National Health Service, or the Department of Health.

This study would not be possible without all the participants who have given their time and support. We thank all the participants and their families. We thank the many research administrators, health-care and social-care professionals who contributed to setting up and delivering the PHOSP-COVID study at all of the 65 NHS trusts/Health boards and 25 research institutions across the UK, as well as those who contributed to setting up and delivering the ISARIC4C study at 305 NHS trusts/ Health boards. We also thank all the supporting staff at the NIHR Clinical Research Network, Health Research Authority, Research Ethics Committee, Department of Health and Social Care, Public Health Scotland, and Public Health England. We thank Kate Holmes at the NIHR Office for Clinical Research Infrastructure (NOCRI) for her support in coordinating the charities group. The PHOSP-COVID industry framework was formed to provide advice and support in commercial discussions, and we thank the Association of the British Pharmaceutical Industry as well NOCRI for coordinating this. We are very grateful to all the charities that have provided insight to the study: Action Pulmonary Fibrosis, Alzheimer’s Research UK, Asthma and Lung UK, British Heart Foundation, Diabetes UK, Cystic Fibrosis Trust, Kidney Research UK, MQ Mental Health, Muscular Dystrophy UK, Stroke Association Blood Cancer UK, McPin Foundations, and Versus Arthritis. We thank the NIHR Leicester Biomedical Research Centre patient and public involvement group and Long Covid Support.

## Ethical Approvals

Identified patients hospitalised during the SARS-COV-2 pandemic were recruited into the International Severe Acute Respiratory and Emerging Infection Consortium World Health Organization Clinical Characterisation Protocol UK (IRAS260007 and IRAS126600). Written informed consent was obtained from all patients. Ethical approval was given by the South Central– Oxford C Research Ethics Committee in England (reference: 13/SC/0149), Scotland A Research Ethics Committee (reference: 20/SS/0028) and World Health Organization Ethics Review Committee (RPC571 and RPC572l; 25 April 2013).

Following hospital discharge patients were recruited to the PHOSP-COVID study for which written consent was obtained and ethical approval given by Leeds West Research Ethics Committee (Ref: *20/YH/0225)*.

## Data sharing

This is an Open Access article under the CC BY 4.0 license.

The ISARIC4C protocol, data sharing and publication policy are available at https://isaric4c.net. ISARIC4C’s Independent Data and Material Access Committee welcomes applications for access to data and materials (https://isaric4c.net).

The PHOSP-COVID protocol, consent form, definition and derivation of clinical characteristics and outcomes, training materials, regulatory documents, information about requests for data access, and other relevant study materials are available online: https://phosp.org/resource/. Access to these materials can be granted by contacting.

All data used in this study is available within ODAP. Data access criteria and information about how to request access is available online: https://phosp.org/resource/. If criteria are met and a request is made, access can be gained by signing the eDRIS user agreement.

## PPI dissemination

Patient and public involvement have been integral to the PHOSP-COVID study and consortium since conception. The PHOSP PPI group is co-chaired by NOCRI (Kate Holmes) and Asthma and Lung UK (Krisnah Poinasamy) with representation of over 10 relevant charities. Members of the ‘Long-COVID Facebook support group’ are closely involved and a Leicester BRC PPI group consisting of people with lived experience of a hospital admission for COVID-19. Patients and public are embedded within the PHOSP-COVID infrastructure including our working groups, core management group, and executive and steering groups. Patients were involved in the development of the clinical research study including the overarching aims, choice of outcomes, consent processes and the structure of the study visits. Patients review all patient facing material. We have recently completed a joint patient and clinician research priority questions exercise hosted by advisors from the James Lind Alliance to ensure co-ownership of the direction of PHOSP-COVID research.

ISARIC4C has a public facing website and twitter account (@CCPUKstudy). We are engaging with print and internet press, television, radio, news, and documentary programme makers. We will explore distribution of findings with Asthma and Lung UK and take advice from NIHR Involve and GenerationR Alliance Young People’s Advisory Groups.

## Author Contributions

**Felicity Liew** has made substantial contributions to this work including: recruitment of participants, acquisition of clinical samples and data, as well as analysis and interpretation of data. They have co-written this manuscript, including all drafting and revisions. They approve the final version to be published and agree to accountability for all aspects of this work.

**Shubha Talwar** has made substantial contributions to acquisition of nasal and plasma antibody data underlying this study. They have reviewed and approved the data underlying this study and supported drafting and revisions of this work. They approve the final version to be published and agree to accountability for all aspects of this work.

**Andy Cross** has made substantial contributions to acquisition of data underlying this study. They have reviewed and approved the data underlying this study and supported drafting and revisions of this work. They approve the final version to be published and agree to accountability for all aspects of this work.

**Brian J. Willett** has made substantial contributions to the analysis of plasma neutralisation and acquisition of data underlying this study. They have reviewed and approved the data presented in this study and have contributed to revisions of the work. They approve the final version to be published and agree to accountability for all aspects of this work.

**Sam Scott** has made substantial contributions to acquisition of data underlying this study. They have reviewed and approved the data and have supported drafting and revisions of this work. They approve the final version to be published and agree to accountability for all aspects of this work.

**Nicola Logan** has made substantial contributions to acquisition of data underlying this study. They have reviewed and approved the data underlying this study and have supported drafting and revisions of this work. They approve the final version to be published and agree to accountability for all aspects of this work.

**Matthew K. Siggins** has made substantial contributions to the analysis and interpretation of data underlying this study. They have reviewed and approved the data presented in this study and supported drafting and revisions for this work. They approve the final version to be published and agree to accountability for all aspects of this work.

**Dawid Swieboda** has made substantial contributions to acquisition of data for this. They have reviewed and approved the data underlying this study and have supported drafting and revisions of this work. They approve the final version to be published and agree to accountability for all aspects of this work.

**Jasmin K. Sidhu** has made substantial contributions to acquisition of data for this work. They have reviewed and approved the data underlying this study and have supported drafting and revisions of this work. They approve the final version to be published and agree to accountability for all aspects of this work.

**Claudia Efstathiou** has made substantial contributions to acquisition of data underlying this study. They have reviewed and approved the data underlying this study and have supported drafting and revisions of this work. They approve the final version to be published and agree to accountability for all aspects of this work.

**Shona C. Moore** has made substantial contributions to implementation of this work and acquisition of data. They have reviewed and approved the data underlying this study and have supported drafting and revisions of this work. They approve the final version to be published and agree to accountability for all aspects of this work.

**Christopher Davis** has made substantial contributions to acquisition of data. They have reviewed and approved the data underlying this study and have supported drafting and revisions of this work. They approve the final version to be published and agree to accountability for all aspects of this work.

**Noura Mohamed** has made substantial contributions to acquisition of clinical samples for this work. They have reviewed and approved the data underlying this study. They approve the final version to be published and agree to accountability for all aspects of this work.

**Jose Nunag** has made substantial contributions to acquisition of clinical samples for this work. They have reviewed and approved the data underlying this study. They approve the final version to be published and agree to accountability for all aspects of this work.

**Clara King** has made substantial contributions to acquisition clinical samples required for this work. They have reviewed and approved the data underlying this study. They approve the final version to be published and agree to accountability for all aspects of this work.

**A.A. Roger Thompson** has made substantial contributions to conception and implementation of this work as well as acquisition of data for this work. They have reviewed and approved the data underlying this study and have supported drafting and revisions of this work. They approve the final version to be published and agree to accountability for all aspects of this work.

**Sarah L. Rowland-Jones** has made substantial contributions to conception and implementation of this work as well as acquisition of data for this work. They have reviewed and approved the data underlying this study and have supported drafting and revisions of this work. They approve the final version to be published and agree to accountability for all aspects of this work.

**Ewen Harrison** has made substantial contributions to study design as well as data access, linkage and analysis. They have had direct access to and have verified clinical data used in this study. They have also supported drafting and revisions of this work. They approve the final version to be published and agree to accountability for all aspects of this work.

**Annemarie B. Docherty** has made substantial contributions to study design as well as data access, linkage and analysis. They have also supported drafting and revisions of this work. They approve the final version to be published and agree to accountability for all aspects of this work.

**Jennifer K. Quint** has made substantial contributions to study design as well as data access, linkage and analysis. They have had direct access to and have verified clinical data used in this study. They have also supported drafting and revisions of this work. They approve the final version to be published and agree to accountability for all aspects of this work.

**James D. Chalmers** has made substantial contributions to conception and design of this work and have supported drafting and revisions of this work. They approve the final version to be published and agree to accountability for all aspects of this work.

**Ling-Pei Ho** has made substantial contributions to conception and design of this work have also supported drafting and revisions of this work. They approve the final version to be published and agree to accountability for all aspects of this work.

**Alexander Horsley** has made substantial contributions to conception and design of this work and have also supported drafting and revisions of this work. They approve the final version to be published and agree to accountability for all aspects of this work.

**Betty Raman** has made substantial contributions to conception and design of this work and have also supported drafting and revisions of this work. They approve the final version to be published and agree to accountability for all aspects of this work.

**Krisnah Poinasamy** has made substantial contributions to conception and design of this work and has supported drafting and revisions of this work. They approve the final version to be published and agree to accountability for all aspects of this work.

**Michael Marks** has made substantial contributions to conception and design of this work and has supported drafting and revisions of this work. They approve the final version to be published and agree to accountability for all aspects of this work.

**Luke Howard** has made substantial contributions to the design of this work and acquisition of clinical samples. They have also supported drafting and revisions of this work. They approve the final version to be published and agree to accountability for all aspects of this work.

**Onn Min Kon** has made substantial contributions to the design of this work and acquisition of clinical samples. They have also supported drafting and revisions of this work. They approve the final version to be published and agree to accountability for all aspects of this work.

**Susanna Dunachie** has made substantial contributions to conception and design of this work and have also supported drafting and revisions of this work. They approve the final version to be published and agree to accountability for all aspects of this work.

**Rachael A. Evans** is the co-lead of PHOSP-COVID and has made substantial contributions to conception and design of this work. They have also supported drafting and revisions of this work. They approve the final version to be published and agree to accountability for all aspects of this work.

**Louise V. Wain** is the co-lead of PHOSP-COVID and has made substantial contributions to conception and design of this work. They have also supported drafting and revisions of this work. They approve the final version to be published and agree to accountability for all aspects of this work.

**Daniel G. Wootton** has made substantial contributions to conception and design of this work as well as acquisition of samples required by this study. They have supported drafting and revisions of this work. They approve the final version to be published and agree to accountability for all aspects of this work.

**J. Kenneth Baillie** obtained funding, is ISARIC4C consortium co-lead, has made substantial contributions to conception and design of this work and has also supported drafting and revisions of this work. They approve the final version to be published and agree to accountability for all aspects of this work.

**Malcolm G. Semple** obtained funding, is ISARIC4C consortium co-lead, sponsor/protocol chief investigator, has made substantial contributions to conception and design of this work and has also supported drafting and revisions of this work. They approve the final version to be published and agree to accountability for all aspects of this work.

**Christopher Brightling** is the chief investigator of PHOSP-COVID and has made substantial contributions to conception and design of this work. They have also supported drafting and revisions of this work. They approve the final version to be published and agree to accountability for all aspects of this work.

**Sara Fontanella** has made substantial contributions to analysis and interpretation of data underlying this study. They have also contributed to drafting and revisions of this work. They approve the final version to be published and agree to accountability for all aspects of this work.

**Thushan I de Silva** has made substantial contributions to conception and design of this work and has also supported drafting and revisions of this work. They approve the final version to be published and agree to accountability for all aspects of this work.

**Antonia Ho** has made substantial contributions to conception and design of this work and has also supported drafting and revisions of this work. They approve the final version to be published and agree to accountability for all aspects of this work.

**Ryan S. Thwaites** has made substantial contributions to acquisition, analysis and interpretation of the data, and has had direct access to and approval of the data underlying this study. They have co-written this manuscript and contributed to drafting and revisions of this work. They approve the final version to be published and agree to accountability for all aspects of this work.

**Lance Turtle** has made substantial contributions to conception and design of this work as well as acquisition and interpretation of data. They have supported drafting and revisions of this manuscript. They approve the final version to be published and agree to accountability for all aspects of this work.

**Peter J.M. Openshaw** obtained funding, is ISARIC4C consortium co-lead, sponsor/protocol chief investigator, and has made substantial contributions to conception and design of this work. They have also made key contributions to interpretation of data and have co-written this manuscript. They approve the final version to be published and agree to accountability for all aspects of this work.

All investigators within ISARIC4C and the PHOSP-COVID consortia have made substantial contributions to the conception or design of this study and/or acquisition of data for this study. The full list of authors within these groups is available in the supplementary materials.

## Declaration of interests

Felicity Liew has no conflicts of interest. Shubha Talwar has no conflicts of interest. Andy Cross has no conflicts of interest. Brian J. Willett has no conflicts of interest. Sam Scott has no conflicts of interest. Nicola Logan has no conflicts of interest. Matthew K. Siggins has no conflicts of interest. Dawid Swieboda has no conflicts of interest. Jasmin K. Sidhu has no conflicts of interest. Claudia Efstathiou has no conflicts of interest. Shona C. Moore has no conflicts of interest. Christopher Davis has no conflicts of interest. Clara King has no conflicts of interest. A.A. Roger Thompson is supported by a British Heart Foundation (BHF) Intermediate Clinical Fellowship FS/18/13/33281. He receives speaker fees and support to attend meetings from Janssen Pharmaceuticals. Sarah L. Rowland-Jones receives support from UKRI for the PHOSP-Covid study. She has grants from UKRI, GCRF, Rosetrees Trust, BHIVA, EDCTP, Globvac. She is on the data safety monitoring board for Bexero trial in HIV+ adults in Kenya. Ewen Harrison has no conflicts of interest. Annemarie B. Docherty has no conflicts of interest. Jennifer K. Quint has no conflicts of interest. James D. Chalmers is the deputy chief editor of ERS. He receives consulting fees from AstraZeneca, Boehringer Ingelheim, Chiesi, GlaxoSmithKline, Insmed, Janssen, Novartis, Pfizer and Zambon. He has grants from AstraZeneca, Boehringer Ingelheim, GlaxoSmithKline, Gilead Sciences, Grifols, Novartis and Insmed. Ling-Pei Ho has no conflicts of interest. Alexander Horsley is Deputy chair of NIHR Translational Research Collaboration (unpaid role). He is currently supported by UK□Research and Innovation. NIHR and NIHR Manchester BRC. Betty Raman receives support from BHF Oxford Centre of Research Excellence, NIHR Oxford BRC and MRC. She receives honoraria from Axcella therapeutics. Krisnah Poinasamy has no conflicts of interest. Susanna J. Dunachie is supported by an NIHR Global Research Professorship (NIHR300791). She is a member of the PITCH Consortium which has received funding from UK Department of Health and Social Care. She receives support from UKRI as part of “Investigation of proven vaccine breakthrough by SARS-CoV-2 variants in established UK healthcare worker cohorts: SIREN consortium & PITCH Plus Pathway” (MR/W02067X/1), with contributions from UKRI/NIHR through the UK Coronavirus Immunology Consortium (UK-CIC), the Huo Family Foundation and The National Institute for Health & Care Research (UKRIDHSC COVID-19 Rapid Response Rolling Call, Grant Reference Number COV19-RECPLAS). S.J.D. is a Scientific Advisor to the Scottish Parliament on COVID-19 for which she receives a fee. SJD also receives support from UKRI, Huo family foundation and NIHR England. Rachael A. Evans is co-lead of PHOSP-COVID and receives funding from UKRI and NIHR for this study. She also receives fees from Astrazenaca / Evidera for consultancy on Long Covid and from Astrazenaca for consultancy on digital health. She has received speaker fees from Boehringer in June 2021.She has held a role as European Respiratory Society Assembly 01.02 Pulmonary Rehabilitation secretary and is on the American Thoracic Society Pulmonary Rehabilitation Assembly programme committee. Louise V. Wain has received support from UKRI, GSK/Asthma + Lung UK and NIHR for this study. She also receives funding from Orion pharma and GSK. She holds contracts with Genentech and AstraZenaca. She has received consulting fees from Galapagos and Boehringer. She is on the data advisory board for Galapagos and is Associate Editor for European Respiratory Journal. Luke Howard has no conflicts of interest. Onn Min Kon has no conflicts of interest. Thushan I de Silva has no conflicts of interest. Antonia Ho has received support from MRC and for the Coronavirus Immunology Consortium (MR/V028448/1) and is a member of NIHR Urgent Public Health Group (June 2020-March 2021). Daniel G. Wootton has no conflicts of interest. Malcolm G. Semple has received support from NIHR UK, MRC UK and Health Protection Research Unit in Emerging & Zoonotic Infections, University of Liverpool. He acts as an independent external and non-remunerated member of Pfizer’s External Data Monitoring Committee for their mRNA vaccine program(s). He is Chair of Infectious Disease Scientific Advisory Board of Integrum Scientific LLC, Greensboro, NC, USA. He is director of MedEx Solutions Ltd and majority owner of MedEx Solutions Ltd and minority owner of Integrum Scientific LLC, Greensboro, NC, USA. His institution has been in receipt of gifts from Chiesi Farmaceutici S.p.A. of Clinical Trial Investigational Medicinal Product without encumbrance and distribution of same to trial sites. He is a non-renumerated member of HMG UK New Emerging Respiratory Virus Threats Advisory Group (NERVTAG) and has previously been a non-renumerated member of SAGE. Christopher Brightling has received funds from MRC, NIHR and Leicester NIHR BRC to support the PHOSP-COVID study. He has received consulting fees and/or grants from GSK, AZ, Genentech, Roche, Novartis, Sanofi, Regeneron, Chiesi, Mologic and 4DPharma. Ryan S. Thwaites has no conflicts of interest. Lance Turtle is supported by grants from MRC, NIHR, Wellcome Trust, FDA and DHSC. He has received consulting fees from MHRA and speak fees from Eisai Ltd. He has a patent pending with ZikaVac. PJMO reports grants from the EU Innovative Medicines Initiative (IMI) 2 Joint Undertaking during the submitted work; grants from UK Medical Research Council, GlaxoSmithKline, Wellcome Trust, EU-IMI, UK, National Institute for Health Research, and UK Research and Innovation-Department for Business, Energy and Industrial Strategy; and personal fees from Pfizer, Janssen, and Seqirus, outside the submitted work.

## Supplementary figure legends

**Figures S1.**
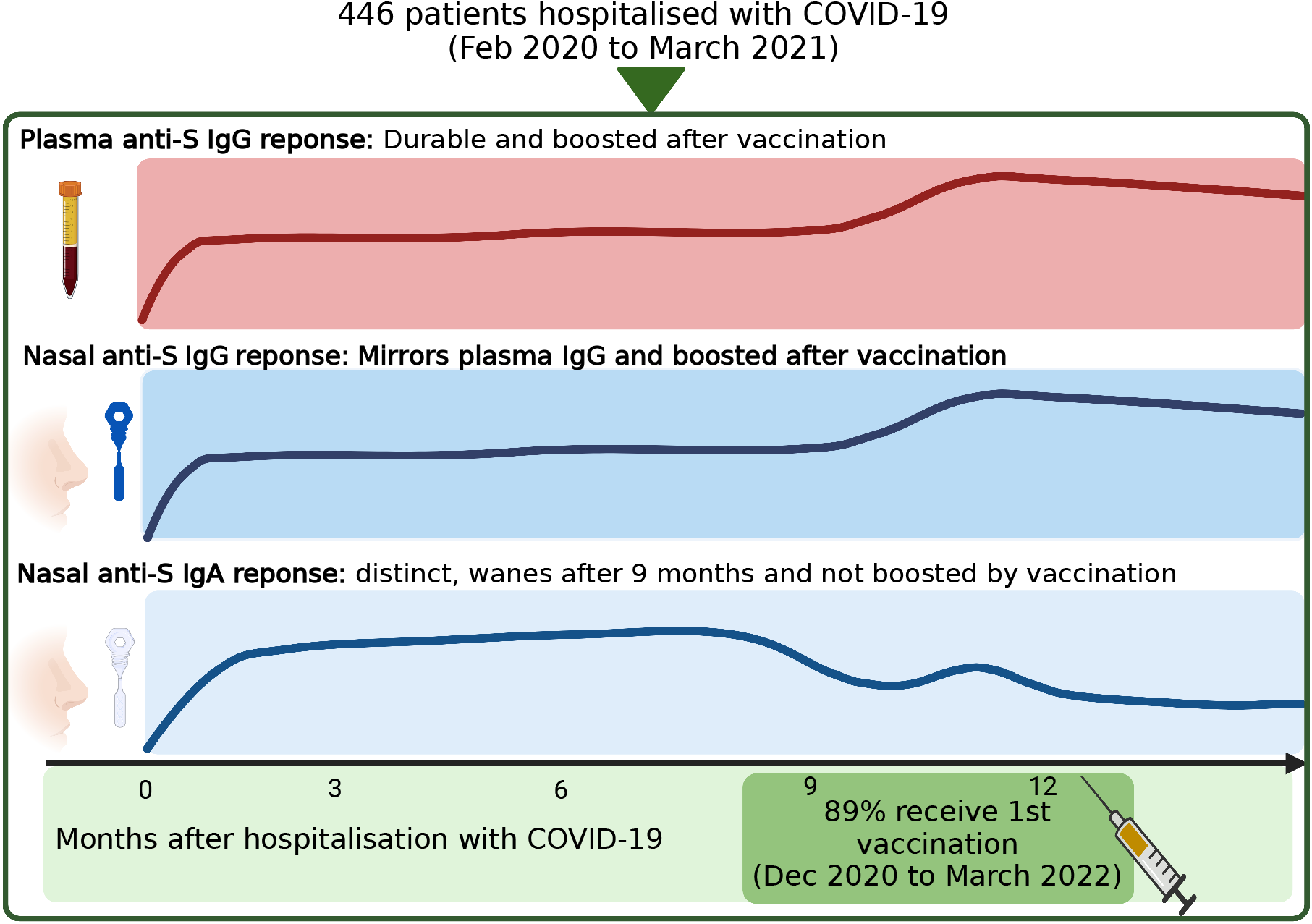
Graphical abstract. Plasma and nasal samples collected at serial intervals from 446 adults hospitalised for COVID-19. Plasma and nasal IgG responses are durable and boosted by vaccination. Nasal IgA responses are compartmentalised from plasma IgG responses and are minimally affected by vaccination. This image was created with BioRender.com

**Figure S2.**
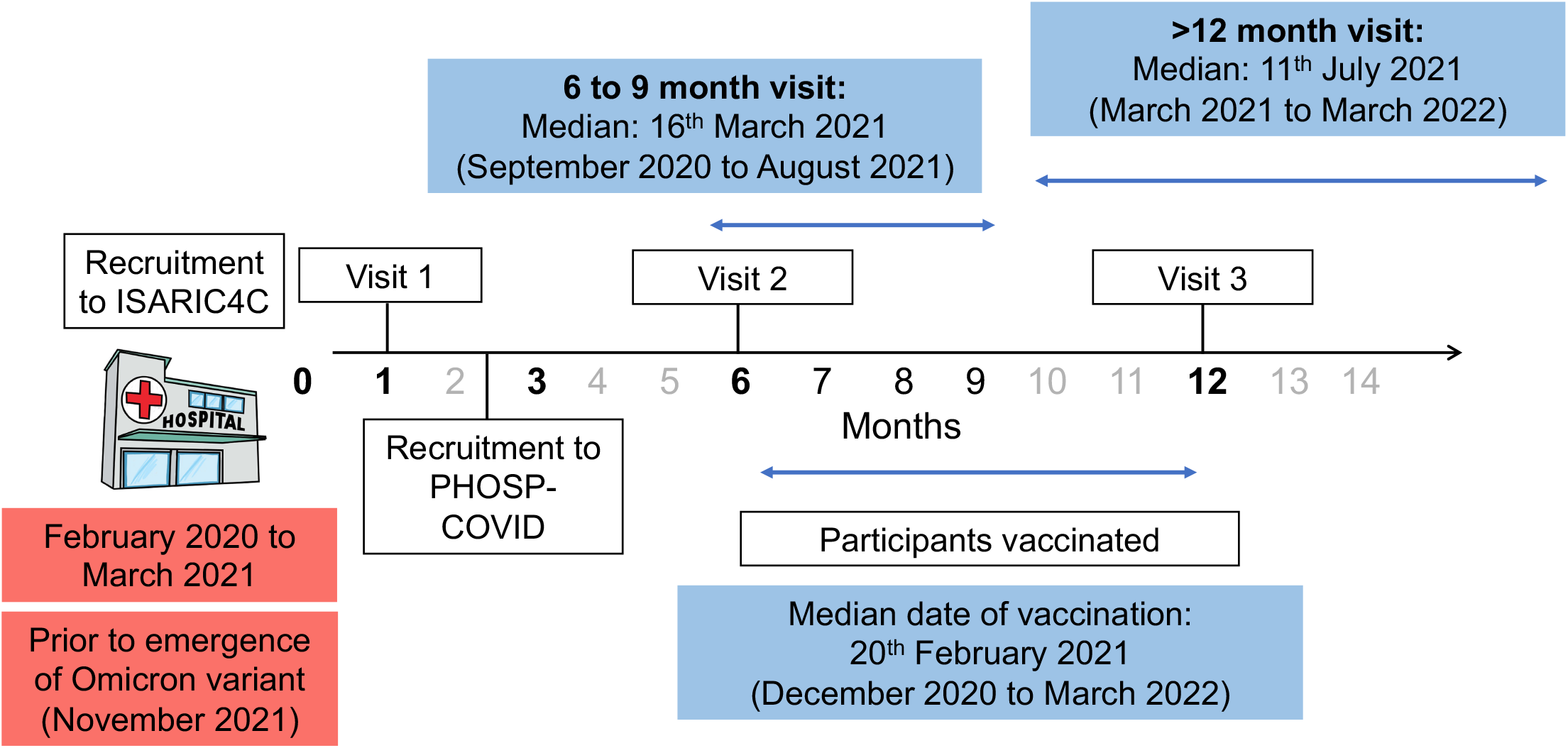
Schematic of study design. Clinical data, plasma and/or nasal samples were obtained during hospital admission and/or 1 to 3 visits during convalescence. The 6 to 9 month visit coincided with the start of the UK vaccination campaign. Vaccination dates are shown as median (range) for individuals where vaccination status was known. Dates in which all study participants attended their 6 to 9 month and >12 month visit are shown in median (range).

**Figure S3.**
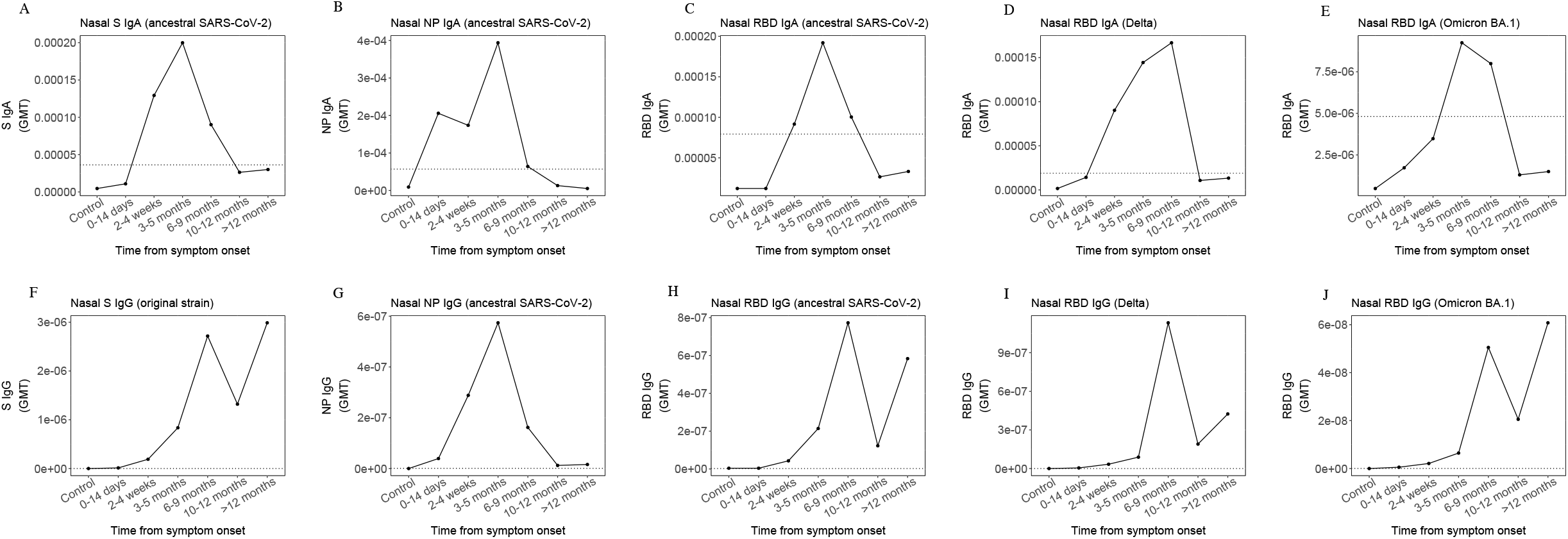
Nasal IgA (A-B) and Nasal IgG (F-G) geometric mean titre (GMT) to S and NP from ancestral SARS-CoV-2. Nasal IgA (C-E) and IgG (G-I) GMT to RBD of ancestral SARS-CoV-2, Delta and Omicron BA.1 variant are also shown. The horizontal dotted line indicates the threshold titre derived from GMT+2SD of pre-pandemic samples.

**Figure S4.**
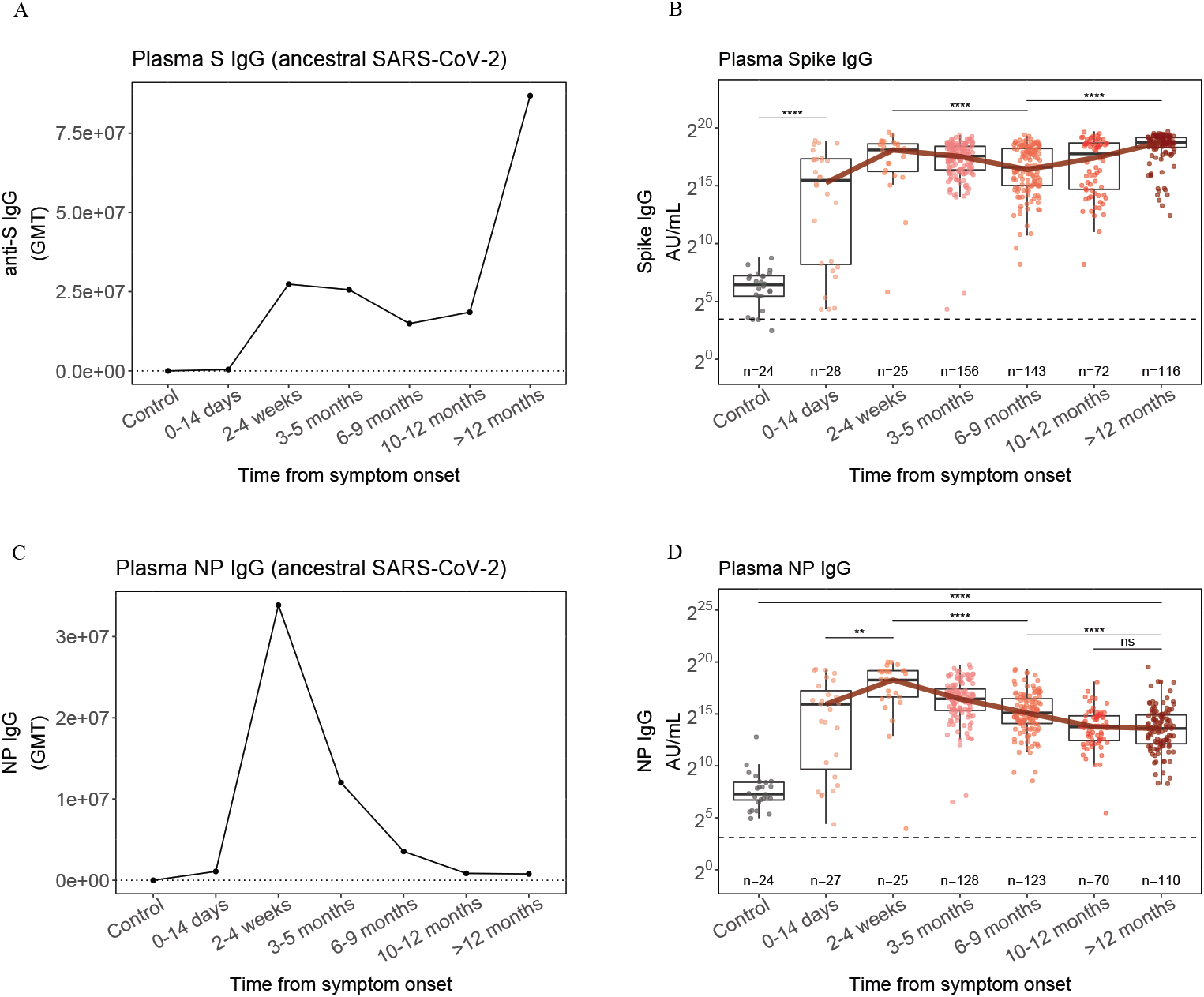
Geometric mean titre (GMT) of plasma anti-S IgG (A) relative to a threshold defined by GMT+2SD of pre-pandemic samples (horizontal dotted line). This has been compared to the WHO threshold titre for seropositivity (horizontal dashed line) (B). The same comparison is shown for anti-NP plasma IgG responses (C–D).

**Figure S5.**
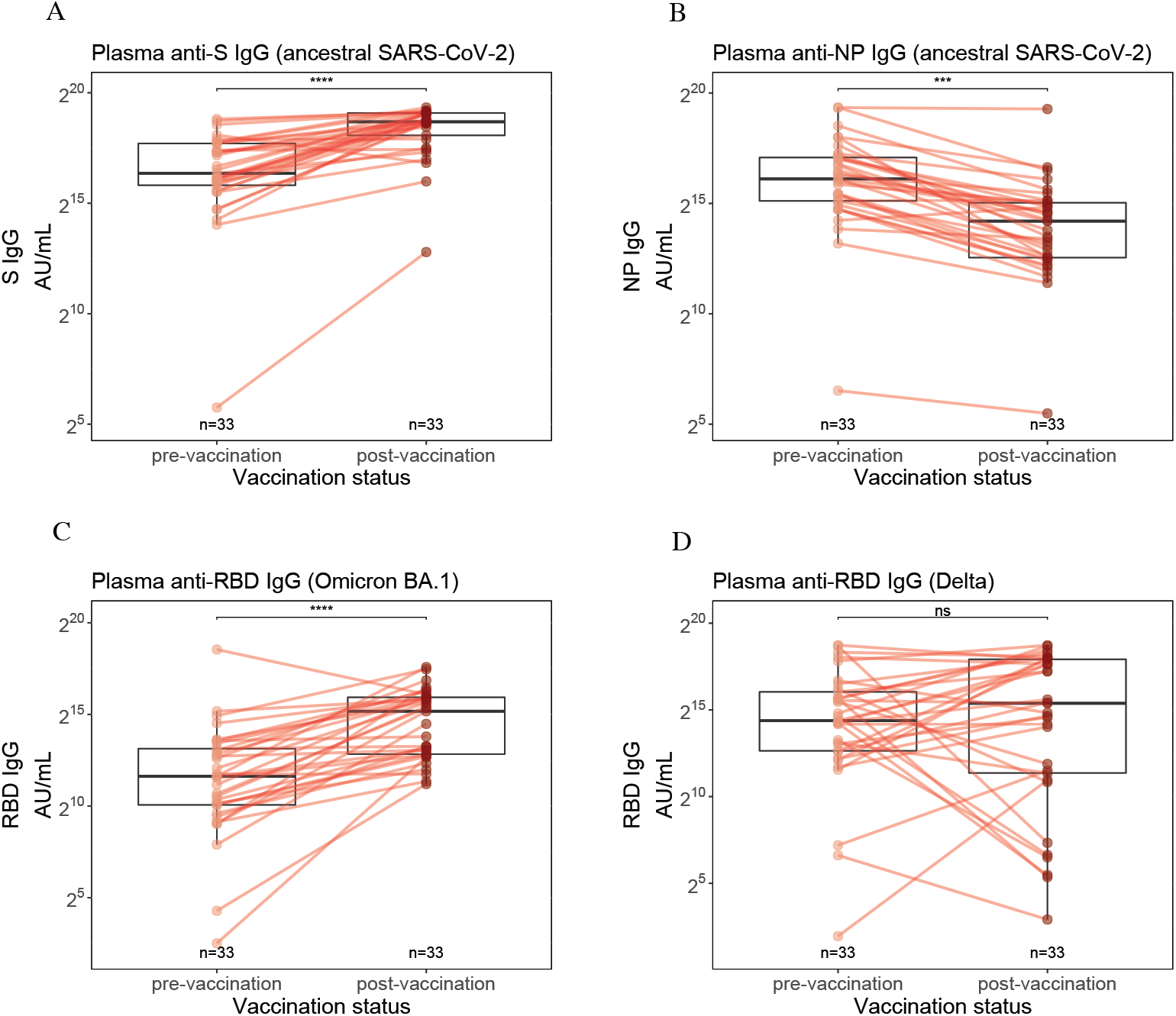
Paired plasma IgG responses to S (A), NP (B) and RBD of Omicron BA.1 (C) and Delta (D) variant, taken before and after vaccination.

**Figure S6.**
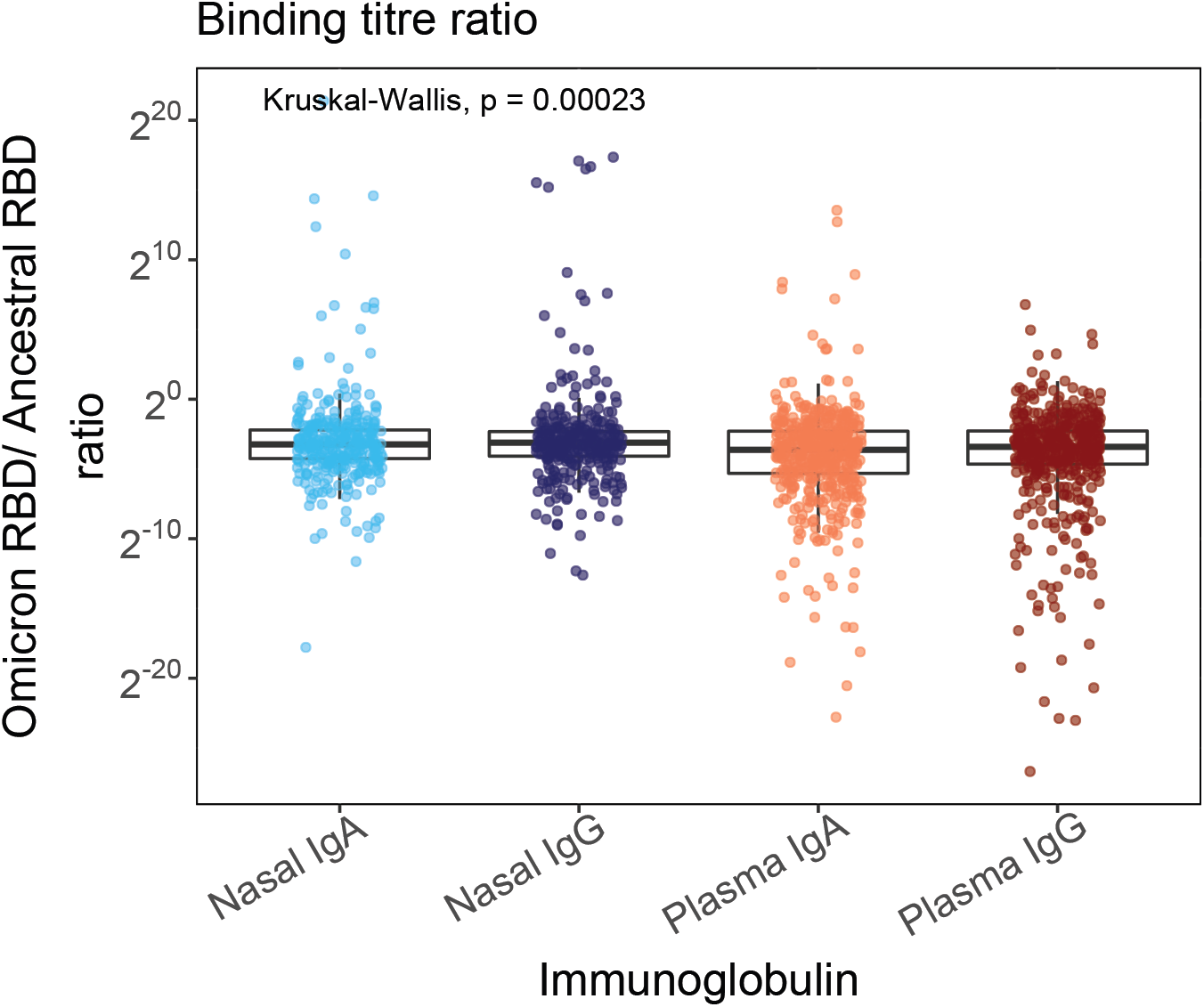
Ratio of binding titre to RBD of Omicron BA.1 variant and ancestral SARS-CoV-2 across nasal and plasma compartments.

**Figure S7.**
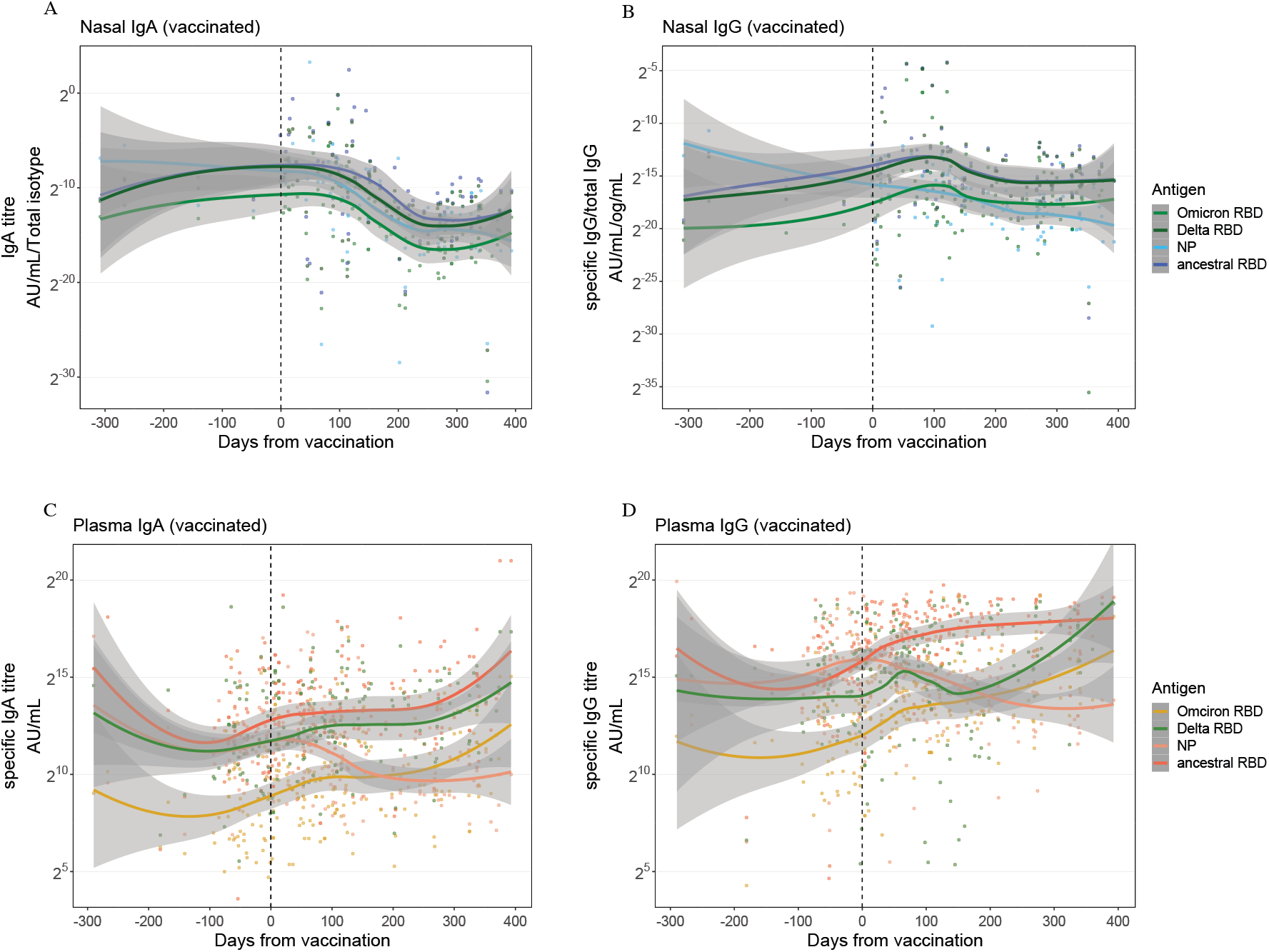
Trajectory of Nasal IgA (A), nasal IgG (B), plasma IgA (C) and plasma IgG (D) responses to RBD of Omicron (BA.1) and Delta variant before and after vaccination. Responses to NP and RBD of ancestral SARS-CoV-2 are also shown for comparison. Trajectories have been modelled using a LOESS regression curve and 95% confidence intervals are shown in grey. The vertical dashed line indicates the time of vaccination.

**Figure S8.**
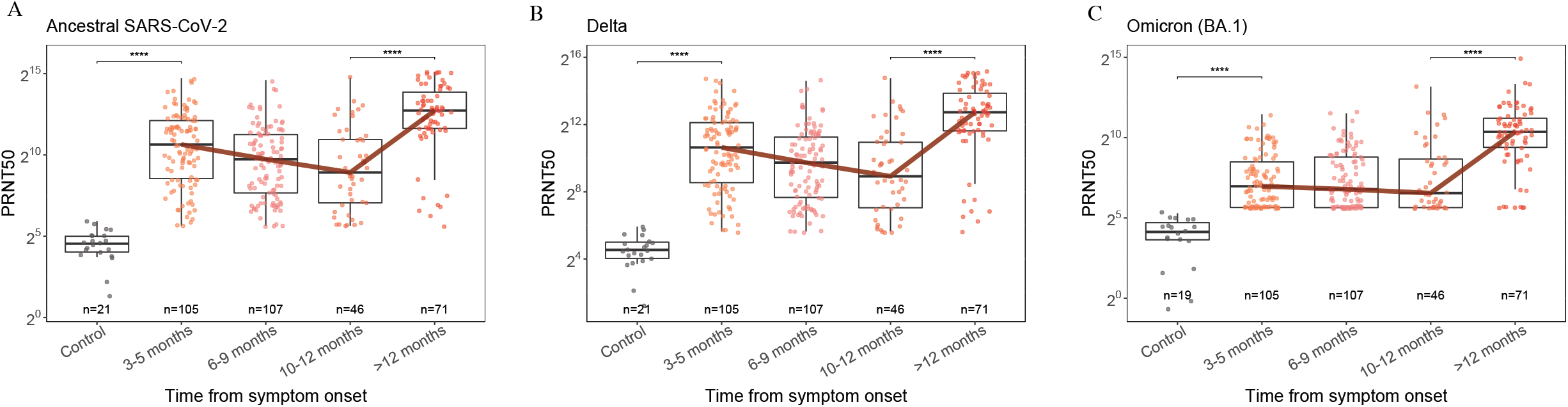
Plasma neutralising titres between 3 and 12 months after infection. Neutralisation of ancestral SARS-CoV-2 (A), Delta variant (B) and Omicron variant (C) are shown. The red line indicates the trajectory of the median titre across each time bin. * = p<0·05, ** = p<0·01, *** = p<0·001, **** = p<0·0001.

**Figure S9.**
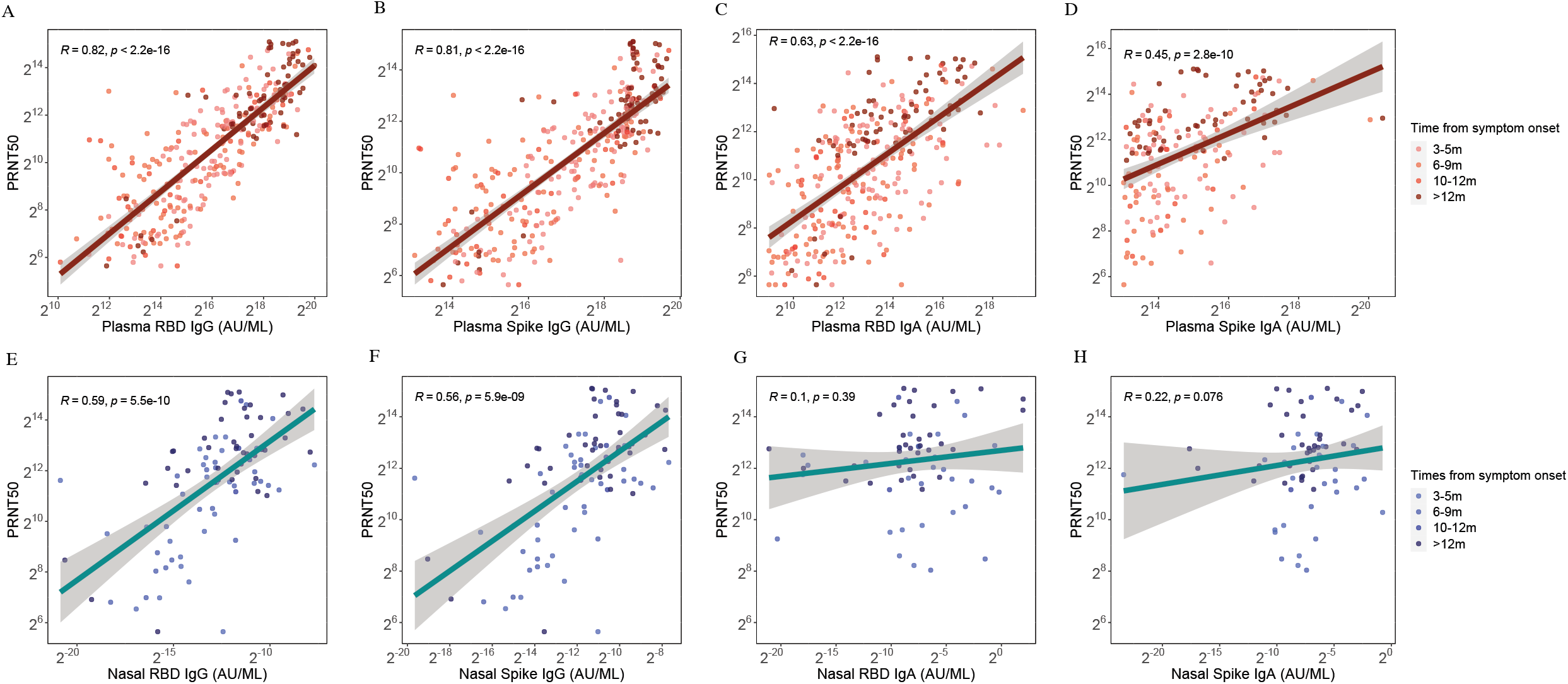
Correlation between plasma neutralising titre and plasma IgG (A–B) and IgA (C–D) binding titre to RBD and S. The correlation between plasma neutralising titre and nasal IgG (E–F) and IgA (G–H) binding titre to RBD and S. A regression line has been fit to the data for which the 95% confidence intervals are shown in grey. *R*=Spearman-rank correlation coefficient. PRNT_50_ = serum dilution resulting in >50% reduction in infectivity.

**Figure S10.**
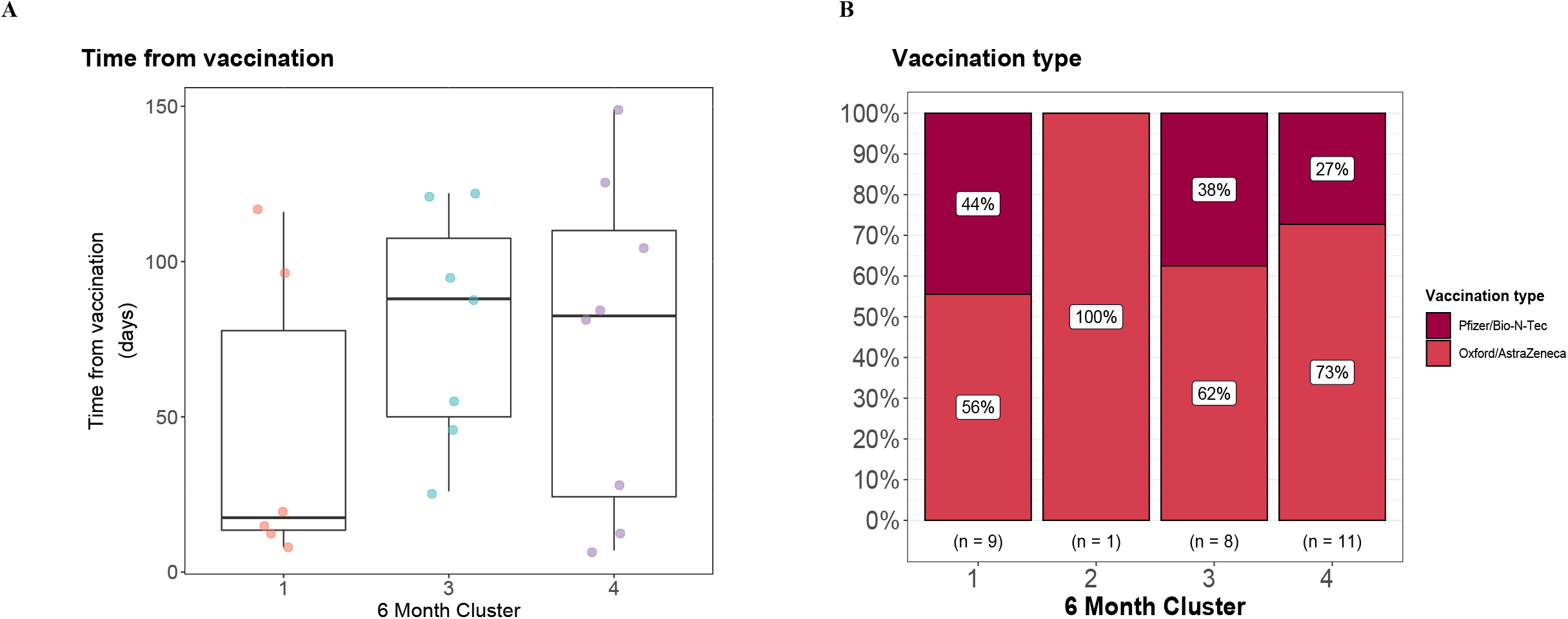
Time from vaccination (A) in cluster 1, 3 and 4 derived from unsupervised, hierarchical clustering analysis of nasal IgA, plasma IgA and plasma IgG anti-S and anti-RBD responses at 6 months from symptom onset. Date of vaccination was not available for any individual in cluster 2. Proportion of individuals vaccinated with either Pfizer/Bio-N-Tec (BNT162b2) or Oxford/ AstraZeneca (ChAdOx1 nCoV-19) vaccine (B) in each cluster.

## Notes

### Competing Interest Statement

A.A. Roger Thompson receives speaker fees and support to attend meetings from Janssen Pharmaceuticals. Sarah L. Rowland-Jones receives support from UKRI GCRF Rosetrees Trust BHIVA EDCTP and Globvac. She is on the data safety monitoring board for Bexero trial in HIV+ adults in Kenya. James D. Chalmers is the deputy chief editor of ERS. He receives consulting fees from AstraZeneca Boehringer Ingelheim Chiesi GlaxoSmithKline Insmed Janssen Novartis Pfizer and Zambon. He has grants from AstraZeneca Boehringer Ingelheim GlaxoSmithKline Gilead Sciences Grifols Novartis and Insmed. Ling-Pei Ho has no conflicts of interest. Alexander Horsley is Deputy chair of NIHR Translational Research Collaboration (unpaid role). He is currently supported by UK□Research and Innovation NIHR and NIHR Manchester BRC. Betty Raman receives support from BHF Oxford Centre of Research Excellence NIHR Oxford BRC and MRC. She receives honoraria from Axcella therapeutics. Krisnah Poinasamy has no conflicts of interest. Susanna J. Dunachie is supported by an NIHR Global Research Professorship (NIHR300791). She is a member of the PITCH Consortium which has received funding from UK Department of Health and Social Care. She receives support from UKRI as part of Investigation of proven vaccine breakthrough by SARS-CoV-2 variants in established UK healthcare worker cohorts: SIREN consortium & PITCH Plus Pathway (MR/W02067X/1), with contributions from UKRI/NIHR through the UK Coronavirus Immunology Consortium (UK-CIC) the Huo Family Foundation and The National Institute for Health & Care Research (UKRIDHSC COVID-19 Rapid Response Rolling Call Grant Reference Number COV19-RECPLAS). S.J.D. is a Scientific Advisor to the Scottish Parliament on COVID-19 for which she receives a fee. SJD also receives support from UKRI Huo family foundation and NIHR England. Rachael A. Evans is co-lead of PHOSP-COVID and receives funding from UKRI and NIHR for this study. She also receives fees from Astrazenaca / Evidera for consultancy on Long Covid and from Astrazenaca for consultancy on digital health. She has received speaker fees from Boehringer and has held a role as European Respiratory Society Assembly 01.02 Pulmonary Rehabilitation secretary and is on the American Thoracic Society Pulmonary Rehabilitation Assembly programme committee. Louise V. Wain has received support from UKRI GSK/Asthma + Lung UK and NIHR for this study. She also receives funding from Orion pharma and GSK. She holds contracts with Genentech and AstraZenaca. She has received consulting fees from Galapagos and Boehringer. She is on the data advisory board for Galapagos and is Associate Editor for European Respiratory Journal. Thushan I de Silva has no conflicts of interest. Antonia Ho has received support from MRC and for the Coronavirus Immunology Consortium (MR/V028448/1) and is a member of NIHR Urgent Public Health Group (June 2020-March 2021). Malcolm G. Semple has received support from NIHR UK MRC UK and Health Protection Research Unit in Emerging & Zoonotic Infections University of Liverpool. He acts as an independent external and non-remunerated member of Pfizer External Data Monitoring Committee for their mRNA vaccine program(s). He is Chair of Infectious Disease Scientific Advisory Board of Integrum Scientific LLC, Greensboro, NC, USA. He is director of MedEx Solutions Ltd and majority owner of MedEx Solutions Ltd and minority owner of Integrum Scientific LLC, Greensboro, NC, USA. His institution has been in receipt of gifts from Chiesi Farmaceutici S.p.A. of Clinical Trial Investigational Medicinal Product without encumbrance and distribution of same to trial sites. He is a non-renumerated member of HMG UK New Emerging Respiratory Virus Threats Advisory Group (NERVTAG) and has previously been a non-renumerated member of SAGE. Christopher Brightling has received funds from MRC, NIHR and Leicester NIHR BRC to support the PHOSP-COVID study. He has received consulting fees and/or grants from GSK, AZ, Genentech, Roche, Novartis, Sanofi, Regeneron, Chiesi, Mologic and 4DPharma. Ryan S. Thwaites has no conflicts of interest. Lance Turtle is supported by grants from MRC, NIHR, Wellcome Trust, FDA and DHSC. He has received consulting fees from MHRA and speak fees from Eisai Ltd. He has a patent pending with ZikaVac. PJMO reports grants from the EU Innovative Medicines Initiative (IMI) 2 Joint Undertaking during the submitted work; grants from UK Medical Research Council, GlaxoSmithKline, Wellcome Trust, EU-IMI, UK, National Institute for Health Research, and UK Research and Innovation-Department for Business, Energy and Industrial Strategy; and personal fees from Pfizer, Janssen, and Seqirus, outside the submitted work.

### Clinical Protocols

https://phosp.org/resource/.

### Author Declarations

This is a combined ISARIC4C and PHOSPCOVID study. Written informed consent was obtained from all patients within ISARIC4C. Ethical approval was given by the South Central Oxford C Research Ethics Committee in England (reference: 13/SC/0149), Scotland A Research Ethics Committee (20/SS/0028) and World Health Organization Ethics Review Committee (RPC571 and RPC572l; 25 April 2013). Written informed consent was obtained from all patients for the PHOSP-COVID study. Ethical approvals for the PHOSPCOVID study were given by Leeds West Research Ethics Committee (20/YH/0225).

