## supplementary materials for "Nasal IgA wanes 9 months after hospitalisation with COVID-19 and is not induced by subsequent vaccination"

**Supplementary methods**

**Recruitment and Ethical approvals**

Identified adults hospitalised during the SARS-COV-2 pandemic were recruited into the International Severe Acute Respiratory and Emerging Infection Consortium (ISARIC) World Health Organization Clinical Characterisation Protocol UK (IRAS260007 and IRAS126600). Written informed consent was obtained from all patients. Ethical approval was given by the South Central–Oxford C Research Ethics Committee in England (reference: 13/SC/0149), Scotland A Research Ethics Committee (20/SS/0028) and World Health Organization Ethics Review Committee (RPC571 and RPC572l; 25 April 2013).

After hospital discharge patients >18 years old who had no co-morbidity resulting in a prognosis of <6 months, were recruited to the PHOSP-COVID study. Written informed consent was obtained from all patients. Ethical approvals for the PHOSP-COVID study were given by Leeds West Research Ethics Committee (20/YH/0225).

Control samples were collected from healthy volunteers without respiratory disease or symptoms of infection prior to the COVID-19 pandemic. Written consent was obtained for all individuals and ethical approvals were given by London- Harrow Research Ethics Committee (13/LO/1899).

**Nasal fluid sampling**

Nasal fluid was collected by placing a NasosorptionTM FX·I device (Hunt Developments UK Ltd) against the inferior turbinate for 1 minute. Nasosorption uses a synthetic absorptive matrix which enables elution of concentrated nasal fluid. The sample was placed immediately on ice and then stored at -80°C. Nasosorption strips were later thawed at room temperature and fluid was eluted using methods previously described, with the addition of 1% (final v/v) Triton-X to elution buffers to inactivate SARS-CoV-2.^1^ Nasal fluid was aliquoted and stored at -80°C.

**Plasma sampling**

Whole blood was collected into a Lithium Heparin Sep tube by venepuncture. Samples were centrifuged at 500g and room temperature for 10 minutes. EDTA plasma was then collected, aliquoted and stored at -80°C.

**Immunoassays**

Antibody responses were measured by MSD electrochemiluminescence multiplex assay (Mesoscale Diagnostics, Rockville, Maryland, USA). Nasal and plasma IgA and IgG responses to Spike (S), Nucleocapsid (NP) and the Receptor-Binding-Domain of Spike (RBD) antigens of ancestral (B.1 lineage) SARS-CoV-2 were measured using MSD V-PLEX COVID-19 Coronavirus Panel 2 Kits, which have been demonstrated to have excellent sensitivity and specificity.^2^ Nasal and plasma antibody responses to RBD antigen of Delta (AY3;AY4;AT4.2;AY5:AY6;AY.7;AY.12;AY.14;B.1.617.2) and Omicron (B.1.1.529; BA.1) variants were measured using MSD V-PLEX SARS-CoV-2 panel 22. The MSD plates consisted of 96 wells each containing 10 pre-coated antigen spots. BSA served as a negative control in each well. Plasma samples were diluted 1 in 5000 and nasal samples 1 in 50 before being analysed on plates according to the manufacturers recommended protocol.^3^ In brief, plates were blocked with MSD blocker A prior to sample analysis to prevent non-specific binding. Diluted samples were incubated followed by addition of MSD SULFO-TAG Anti-Human IgA or IgG antibody to detect bound immunoglobulin. Plates were subsequently measured on a MESO QuickPlex SQ 120 Reader (MSD). An equivalent assay for responses to RBD from ancestral (B.1 lineage) SARS-CoV-2 was present on panel 2 and panel 22 to ensure comparable performance between kits. All values at or below the lower limit of detection (LLOD) were replaced with LLOD. All values at or above the upper limit of detection (ULOD) were replaced with ULOD.

Total IgA and IgG content of nasal fluid was measured using a human antibody isotyping Procarta-Plex protein quantitation immunoassay (Invitrogen, Massachusetts, United States). Nasal samples were diluted 1 in 50 prior to incubation on plates. Plates were prepared and analysed according to the manufacturers protocol (Publication Number MAN0024721).^4^ Plates were read on a BioPlex 200 instrument (MSD). All values at or below the lower limit of detection (LLOD) were replaced with LLOD. All values at or above the upper limit of detection (ULOD) were replaced with ULOD.

**Measurement of virus neutralising Antibodies**

Unless otherwise stated, all cell culture media and supplements were obtained from Thermo Fisher Scientific, Paisley, UK. HEK293, HEK293T, and 293-ACE2 cells were maintained in Dulbecco’s modified Eagle’s medium (DMEM) supplemented with 10% foetal bovine serum, 200mM L-glutamine, 100µg/ml streptomycin and 100 IU/ml penicillin. HEK293T cells were transfected with the appropriate SARS-CoV-2 S gene expression vector (wild type or variant) in conjunction with p8.91^8^ and pCSFLW^9^ using polyethylenimine (PEI, Polysciences, Warrington, USA). HIV (SARS-CoV-2) pseudotypes containing supernatants were harvested 48 hours post-transfection, filtered at 0.45mm, aliquoted, and frozen at -80^o^C prior to use. The SARS-CoV-2 spike glycoprotein expression constructs were synthesised by GenScript (Netherlands). Constructs bore the following mutations relative to the Wuhan-Hu-1 sequence (GenBank: MN908947): **B.1 (Wuhan D614G)** – D614G; **B.1.1.7 (Alpha)** - Δ69-70, Δ144, N501Y, A570D, D614G, P681H, T716I, S982A, D1118H; **B.1.617.2 (Delta)** – T19R, G142D, Δ156-157, R158G, L452R, T478K, D614G, P681R, D950N. **B.1.1.529 (Omicron BA.1) -** A67V, Δ69-70, T95I, G142D/Δ143-145, Δ211/L212I, ins214EPE, G339D, S371L, S373P, S375F, K417N, N440K, G446S, S477N, T478K, E484A, Q493R, G496S, Q498R, N501Y, Y505H, T547K, D614G, H655Y, N679K, P681H, N764K, D796Y, N856K, Q954H, N969K, L981F**.** All synthesised S genes were codon-optimised, incorporated the mutation K1255STOP to enhance surface expression, and were cloned into the pcDNA3.1(+) eukaryotic expression vector. 293-ACE2 target cells^10^ were maintained in complete DMEM supplemented with 2µg/ml puromycin.

Neutralising activity in each sample was measured by a serial dilution approach. Each sample was serially diluted in triplicate from 1:50 to 1:36450 in complete DMEM prior to incubation with HIV (SARS-CoV-2) pseudotypes, incubated for 1 hour, and plated onto 239-ACE2 target cells. After 48-72 hours, luciferase activity was quantified by the addition of Steadylite Plus chemiluminescence substrate and analysis on a Perkin Elmer EnSight multimode plate reader (Perkin Elmer, Beaconsfield, UK). Antibody titre was then estimated by interpolating the point at which infectivity had been reduced to 90% of the value for the no serum control samples.

**Sample size calculations**

To assess if the sample size was sufficient to analyse the effect of time, disease severity and vaccination on median antibody titre, we performed sample size calculations based on previously published data. Calculations (shown below) demonstrated that sample size was sufficient for the planned analyses. Calculations were generated using G*Power version 3.1.

From published data on antibody titres between time-points^5^:

Two-tailed Kruskal Wallis/ Anova

Effect size = 0.66

Sample size to achieve power 0.8 (type-1 error 0.05) = **20**

From published data on the effect of vaccination on antibody titres after SARS-CoV-2 infection^6^:

Two-tailed Wilcoxon signed rank test (paired)

Effect size = 1.24

Sample size to achieve power 0.8 (type-1 error 0.05) = **18**

From published data on the influence of disease severity on serum antibody responses to SARS-CoV-2^7^ :

Two-tailed Wilcoxon-signed rank test (unpaired)

Effect size = 1.36

Sample size to achieve power 0.8 (type-1 error 0.05) = **20**

**Data analysis**

Analyses were conducted within the National Safe Haven using the Outbreak Data Analysis Platform (ODAP). Statistical analyses used R version 4.2.0. All tests were two-tailed and statistical significance was defined as a *p*-value<0.05 after adjustment for false discovery rate (q-value=0.05).

The data were confirmed to be non-parametrically distributed using quantile Vs quantile plots. To understand the durability of antibody responses, samples were grouped into time-bins according to the distribution of data over time from symptom onset and to enable statistical comparison between time points and controls. Where date of symptom onset was missing, time from symptom onset was approximated according to the visit at which the sample was collected and the date of admission, if known. Comparisons between timepoints were made using the optimal pooled t-test, which performs well in non-parametric partially paired data.^8^

To explore the effect of vaccination on the antibody response, a LOESS regression curve was fitted to data from repeated and cross-sectional samples taken before and/or after vaccination, from those who were known to be vaccinated. The anti-NP and anti-S trajectories before and after vaccination were compared visually using the LOESS regression.

To explore the relationship between plasma and nasal responses, as well as their relationship to age and disease severity, variables were analysed in a correlation matrix, measuring the Spearman rank correlation coefficient between variables. All variables were scaled and centred prior to analysis. The variables in the correlogram were hierarchically clustered using Ward’s minimum variance, minimising the Euclidian distance between variables. The analysis was undertaken and visualised using the ‘corrplot’ package in R version 4.0.5.

To further explore the factors determining the relationship between nasal IgA and plasma responses after vaccination, unsupervised clustering was performed using hierarchical clustering with Ward’s minimum variance, comparing the Euclidian distance between individuals who had samples taken at 6 to 9 months. For any paired or repeated measures within the time frame, the latter of the two time points was selected for analysis. All log-transformed antibody variables were scaled and centred prior to analysis. Rows of the heatmap were subsequently annotated with individual’s WHO clinical progression score and age to determine if either factor associated with clusters. The number of clusters was determined using the Silhouette score. To understand how vaccination might affect cluster membership, the mean time from vaccination was compared between each cluster using the Kruskal-wallis test. The proportion of individuals in each cluster receiving ChAdOx1 nCoV-19 vaccine was compared using the chi-squared test. The analyses were undertaken and visualised using the ‘cluster’, ‘ggplot2’, ‘ggstatsplot’, ‘factoextra’ and ‘pheatmap’ package in R version 4.0.5.

**Role of the funding source**

ISARIC4C is supported by grants from the National Institute for Health and Care Research (award CO-CIN-01) and the Medical Research Council (grant MC_PC_19059) Liverpool Experimental Cancer Medicine Centre provided infrastructure support for this research (grant reference: C18616/A25153). The PHOSP-COVD study is jointly funded by UK Research and Innovation and National Institute of Health and Care Research (grant references: MR/V027859/1 and COV0319). The funders were not involved in the study design, interpretation of data or the writing of this manuscript. See ‘Acknowledgements’ for the full list of grants which have contributed to this work.
